# Dysbiosis in metabolic genes of the gut microbiomes of patients with an ileo-anal pouch resembles that observed in Crohn’s Disease

**DOI:** 10.1101/2020.09.23.20199315

**Authors:** Vadim Dubinsky, Leah Reshef, Keren Rabinowitz, Karin Yadgar, Lihi Godny, Keren Zonensain, Nir Wasserberg, Iris Dotan, Uri Gophna

**Affiliations:** The Shmunis School of Biomedicine and Cancer Research, George S. Wise Faculty of Life Sciences, Tel-Aviv University, Tel Aviv, Israel; The Division of Gastroenterology, Rabin Medical Center, Petah-Tikva, Israel; Felsenstein Medical Research Center, Rabin Medical Center, Petah Tikva, Israel; Sackler Faculty of Medicine, Tel-Aviv University, Tel Aviv, Israel; Colorectal Unit, the Division of Surgery, Rabin Medical Center, Petah-Tikva, Israel

**Keywords:** Pouchitis, UC, CD, mucin, butyrate, bile acids, oxidative stress, classifier

## Abstract

**Background:** Crohn’s disease (CD), ulcerative colitis (UC) and pouchitis are multifactorial and chronic inflammatory diseases of the gastrointestinal tract, termed together as inflammatory bowel diseases (IBD). Pouchitis develops in former patients with UC after total proctocolectomy and ileal pouch-anal anastomosis (“pouch surgery”) and is characterized by inflammation of the previously normal small intestine comprising the pouch. It has been extensively shown that broad taxonomic and functional alterations (“dysbiosis”) occur in the gut microbiome of patients with IBD. However, the extent to which microbial dysbiosis in pouchitis resembles that of CD or UC has not been investigated in-depth, and the pathogenesis of pouchitis largely remains unknown.

**Results:** In this study we collected 250 fecal metagenomes from 75 patients with a pouch, including both non-inflamed (normal pouch) and inflamed (pouchitis) phenotypes, and compared them to publicly available metagenomes of patients with CD (n=88), and UC (n=76), as well as healthy controls (n=56). Patients with pouchitis presented the highest level of dysbiosis compared to other IBD phenotypes based on species, metabolic pathways and enzyme profiles, and their level of dysbiosis was correlated with intestinal inflammation. In patients with pouchitis, the microbiome mucin degradation potential was lower, but was accompanied by an enrichment of *Ruminococcus gnavus* strains encoding specific mucin-degrading glycoside hydrolases, which might be pro-inflammatory. Butyrate and secondary bile acids producers were decreased in all IBD phenotypes and were especially low in pouchitis. Butyrate synthesis genes were positively correlated with total dietary fiber intake. Patients with pouchitis harbored more facultative anaerobic bacteria encoding enzymes involved in oxidative stress response, suggesting high oxidative stress during pouch inflammation. Finally, we have developed enzymes-based classifiers that can distinguish between patients with a normal pouch and pouchitis with an area under the curve of 0.91.

**Conclusions:** We propose that the non-inflamed pouch is already dysbiotic (function- and species-wise) and microbially is more similar to CD than to UC. Our study reveals microbial functions that underlie the pathogenesis of pouchitis and suggests bacterial groups and functions that could be targeted for nutritional intervention to attenuate or prevent small intestinal inflammation present in pouchitis and CD.

## Background

Inflammatory bowel diseases (IBD), including Crohn’s disease (CD), ulcerative colitis (UC) and pouchitis, are chronic, relapsing and remitting inflammatory conditions of the intestine. CD mainly affects the small intestine and colon, while UC is localized in the colon. The etiology of IBD is multifactorial, involving genetic predisposition [1], environmental triggers [2], and an aberrant immune-mediated response towards specific antigens of the intestinal microbiota [3].

Approximately 25% of patients with complicated UC may undergo total large bowel resection. Reconstruction of intestinal continuity is achieved by the creation of a reservoir (“pouch”) from the healthy small intestine which is connected to the anus (ileal pouch-anal anastomosis, “pouch surgery”) [4]. Up to 60% of these former UC patients may develop inflammation of the previously normal small intestine comprising the pouch (pouchitis) [5]. Pouchitis is the most common complication after pouch surgery, but its pathogenesis is still largely unknown [6].

It has been previously shown that the gut microbiome in patients with IBD is characterized by lower diversity, lower stability and compositional changes in specific taxa compared to healthy individuals [7]. Levels of facultative anaerobes from *Enterobacteriaceae* are increased while beneficial obligate anaerobes such as *Faecalibacterium prausnitzii* and *Eubacterium rectale* are decreased [8, 9, 10, 11, 12]. Shifts in microbial functions in the metagenomes of patients with IBD had also been demonstrated, e.g. lower presence of short-chain fatty acid (SCFA) synthesis pathways, increase in oxidative stress response genes, and alteration in amino-acids metabolism genes [9, 13]. These taxonomic and functional changes are broadly referred to as “dysbiosis”.

We had previously demonstrated that patients with a pouch and CD might have several mechanisms of pathogenesis in common including mucosal gene and microRNA expression [14, 15], anti-glycan serologic responses [16], and CD-like bacterial dysbiosis [11]. Yet, the extent to which microbial dysbiosis in pouchitis resembles that of CD has not been investigated in depth. Here we analyzed the shotgun metagenomes of patients with a pouch, IBD and healthy subjects with the following aims: 1) To identify microbial signatures (species and functional genes) that might be common to disease phenotypes and thus to place patients with a pouch in the context of “primary” IBD. More specifically, we wanted to test whether pouchitis has microbial features that are more similar to those of CD. 2) To examine several key bacterial functions that may underlie the pathogenesis of pouchitis: excessive mucin degradation, reduced butyrate and secondary bile acids production, and increased bacterial resistance to oxidative stress.

## Methods

### Study cohorts description and metadata characteristics

Patients after pouch surgery were recruited at a dedicated pouch clinic in a tertiary IBD referral center in Israel (Rabin Medical Center [RMC]). The study was approved by the local institutional review board (0298–17) and the National Institutes of Health (NCT01266538). Demographic and clinical data were obtained during clinic visits. After signing informed consent, patients were followed prospectively, and clinic visits including fecal sample collection were scheduled every 3 months or when patients had an episode of pouchitis. Fecal samples were collected in sterile cups and immediately frozen at −80°C until processing. Fecal calprotectin levels were measured. Pouch disease behavior (phenotype) was defined as normal pouch, acute/recurrent-acute pouchitis, chronic pouchitis or Crohn’s like disease of the pouch (CLDP); see Supplementary Materials and Methods for a detailed description. Disease activity was defined based on global physician assessment and on the clinical subscore of the pouchitis disease activity index [^17^].

For the exploratory analysis done in this study, the patients were divided into two groups (discovery cohort): normal pouch (n=35, 103 fecal samples) and pouchitis (n=34, 105 fecal samples). Normal pouch group included samples from patients with a normal pouch (with UC or familial adenomatous polyposis [FAP] background), while the pouchitis group included samples from patients with chronic pouchitis and CLDP. Samples from patients with recurrent-acute pouchitis phenotype were set aside for testing the pouchitis prediction classifier (validation cohort; see Machine learning classifiers part). For these patients, the clinical phenotype defined during the last follow-up clinic visit was recorded. In the validation cohort, 42 samples from 19 patients were included and owing to the longitudinal nature of the cohort, 13 of these patients had also a sample in the discovery cohort which were treated independently. For dietary fiber information obtained for a subset of 52 samples, see Supplementary Materials and Methods.

We excluded samples collected during or within 1-month of antibiotics treatment. 84% of the pouch samples in our dataset were off-antibiotics for 180 days or longer and the rest were off-antibiotics between 1 to 6-months (Supplementary Table 1A, B). While patients with pouchitis tend to have a history of high antibiotic use, after 1-month off antibiotics, the microbiome diversity is significantly increased, and following 6-months post-antibiotics it recovers to pre-treatment levels [12]. Nevertheless, to account for any remaining confounding antibiotics effects, we also included antibiotic use (1-month or 6-months without antibiotics) as a covariate in the models.

**Table 1.**
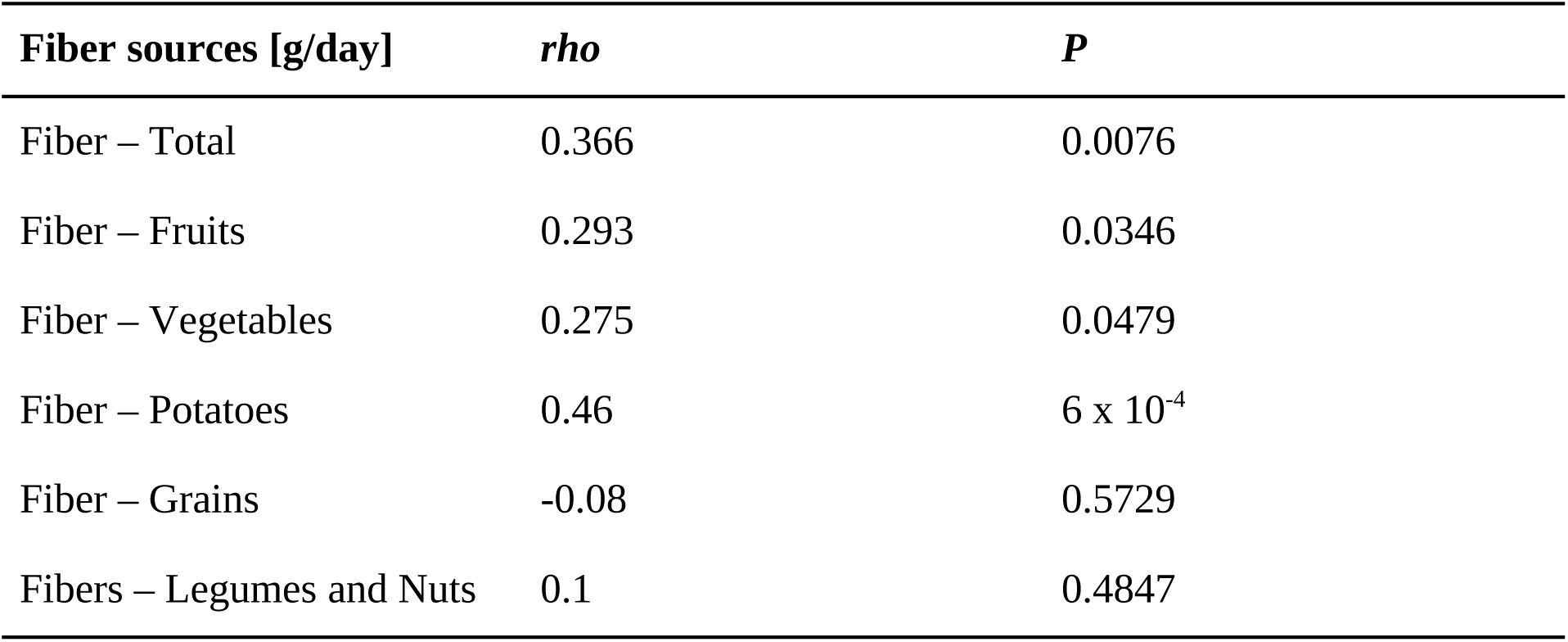
Spearman correlation between dietary fiber intake and the abundance of butyrate genes (*but* +*buk*) in patients with a pouch samples [n=52].

Non-pouch IBD (patients with UC and CD) and healthy adult metagenomes were obtained from publicly available data, previously published in [18], and are part of the PRISM, LifeLines DEEP and NLIBD cohorts. We analyzed these metagenomes with the same bioinformatic pipeline as the pouch metagenomes as described in the following sections, starting the analysis from raw reads.

### DNA Extraction and Shotgun Metagenomic Sequencing of Pouch Samples

DNA was extracted from fecal samples as described in [11]. Genomic libraries were prepared with the Nextera XT library preparation kit using approximately 1 ng of total DNA per sample (DNA concentrations verified by Qubit fluorometry). Metagenomic sequencing was done using Illumina (San Diego, CA) NextSeq500 paired-end (2 x 150 base pair reads) at DNA Services Facility, University of Illinois, Chicago, IL. Sequencing reads were quality filtered with Trimmomatic v0.36 [19] using default parameters, thus removing low-quality and short reads, low-quality bases, and clipping Illumina adaptors. Human host DNA reads were removed by mapping metagenomic reads against the GRCh38 human genome (build 38) with Bowtie2 version-2.2.9 [20] in *--very-sensitive-local* mode. After reads trimming and human reads removal, the mean sequence depth (± standard deviation) was 0.93 ± 0.3 Gbp (6.22 ± 2 million reads) per sample.

### Taxonomic and functional profiling of the metagenomic datasets

Taxonomic profiling was performed using MetaPhlAn2 classifier v2.6.0 [21] which classifies metagenomic reads by mapping to a database of clade-specific marker genes. MetaPhlAn2 was run with the following parameters changed from default: --tax_lev set to “s” (classify taxonomy to species level), --ignore_virus, and --ignore_eukaryotes to ignore viral and eukaryote reads, respectively. Species relative abundance tables from all samples were merged. Species present in less than 5% of the samples and at less than 0.1% relative abundance in any individual sample were removed. For all downstream statistical analysis, we used the bacterial species that are common to the pouch and non-pouch metagenomes (species intersection). This procedure can minimize potential confounders such as species that appear solely in one dataset due to country-specific differences. The only exception was for the Shannon diversity index calculation, where all taxa (union) from both datasets were used in order to obtain a more complete and accurate diversity trend.

Functional profiling was performed with HUMAnN2 v0.11.1 [22]. Briefly, HUMAnN2 first maps metagenomic reads (nucleotide-level with Bowtie2) against functionally annotated pangenomes of the species identified during taxonomic profiling. Reads that do not map to any identified pangenome are subjected to a translated search with DIAMOND [23] against the UniRef90 protein database, and hits are weighted according to alignment quality, sequence length, and coverage. Gene-level outputs are produced in reads per kilobase units and stratified according to known/unclassified microbiome contributions. Per-sample gene abundances were total-sum scaled normalized (to account for the number of reads in each metagenome) to copies per million (cpm) units. The obtained gene abundances were regrouped to MetaCyc metabolic pathways and enzymes (Enzyme Commission number [EC]).

### Screening the metagenomes for key bacterial functions with custom curated databases

For the identification and quantification of butyrate synthesis pathways, an extensive database from [24] was used, which encompassed hidden Markov models (HMM) on full length proteins considering the entire taxonomic diversity of butyrate producers that contained 1,716 genomes. A modified version of this database was used so that only four terminal enzymes in the main pathways known for butyrate production from each of the genomes were considered: in both acetyl-CoA and glutarate pathways, the terminal enzymes are butyryl-CoA:acetate CoA transferase (*but*) and butyrate kinase (*buk*). In 4-aminobutyrate pathway, the terminal enzyme is butyryl-CoA:4-hydroxybutyrate CoA transferase (*4hbt*), and in lysine pathway, the terminal enzyme is butyryl-CoA:acetoacetate CoA transferase (*ato*). For the analysis of microbiome-derived secondary bile acids, we used the database from [25], which was built on the full length HMM for the *bai* genes cluster (*baiA-I* operon) that consisted of *baiA, baiB, baiCD, baiE, baiF, baiG, baiH*, baiI genes, obtained from all genomes available in PATRIC.

Quality-filtered metagenomic reads were used as a query in a translated nucleotide search (BLASTX using DIAMOND v0.9.24.125 [23]) against the databases described above. Only the top hit per read was reported (*--max-target-seqs* 1) and the minimum e-value threshold was set to 1 x 10^−5^ (*--evalue*). For butyrate synthesis genes, only reads with sequence similarity matches of ≥ 90% to a reference, and ≥ 25 amino acids alignment length were retained and counted. For bacterial secondary bile acid production genes, the sequence similarity cutoff was lowered to ≥ 70% due to a less comprehensive reference database. Read counts for each of the key functions analyzed (butyrate and secondary bile acids genes) were normalized to RPKM units (Reads per kilobase per million mapped reads), i.e. normalized according to the corresponding median gene length and total-sum scaled according to the number of reads in each metagenome. The key functional genes were assigned to bacterial taxa according to the taxonomic identity of the respective gene entries in the database.

### Mucin degradation glycoside hydrolases (GH) enzymes analysis

For the analysis of glycoside hydrolases (GH) enzymes that are involved in mucin degradation, only GH that were functionally characterized by *in-vitro* activity or transcriptomic assays [26] were considered. Thus the following enzymes from HUMAnN2 EC4 annotations were considered as mucin-related GH: Protein O-GlcNAcase (EC 3.2.1.169), Exo-alpha-sialidase (EC 3.2.1.18), Beta-galactosidase (EC 3.2.1.23), Alpha-N-acetylgalactosaminidase (EC 3.2.1.49), Alpha-N-acetylglucosaminidase (EC 3.2.1.50), Alpha-L-fucosidase (EC 3.2.1.51) and Beta-N-acetylhexosaminidase (EC 3.2.1.52).

To analyze the specific GH of *Ruminococcus gnavus*, GH29 (Alpha-L-fucosidase), GH95 (glycoside hydrolase family 95) and GH33 (Exo-alpha-sialidase) protein sequences of *Ruminococcus gnavus* ATCC 29149 strain JCM6515 were downloaded from Carbohydrate-Active enZYmes (CAZY) database [27]. Three single-copy housekeeping genes (DNA gyrase subunit A [*gyrA*], recombinase A [*recA*], and 50S ribosomal protein L2 [*rplB*]) of the same *R. gnavus* strain were downloaded as well. These GH and the housekeeping genes were used to build a reference database for similarity search as mentioned previously. For GH, sequence similarity matches of ≥ 90% and ≥ 25 amino acids alignment length were used, and for housekeeping genes, sequence similarity matches were increased to ≥ 97% to count only closely related species. Read counts were normalized by the corresponding gene length, and the percentage of *R. gnavus* strains encoding GH29, GH95 and GH33 were calculated by dividing each GH by the mean abundance of *gyrA, recA*, and *rplB*. As *gyrA* recruited the highest number of reads even after gene-length normalization, we used only *gyrA* for the final calculation. Samples with fewer than 5 *gyrA* reads of *R. gnavus* were removed from the analysis.

### Statistical analysis

The Kruskal-Wallis H-test with Dunn’s test for multiple comparisons correction (pairwise tests between phenotypes) was performed using *kruskal*.*test* and *dunn*.*test* functions in R package ‘dunn.test’. Alpha-diversity (within-sample) was calculated with the Shannon diversity index using *diversity* function in R package ‘vegan’. Beta-diversity (between-sample) was calculated with Bray-Curtis dissimilarity (*vegdist* function in R package ‘vegan’) and visualized with principal coordinate analysis *cmdscale* function (classical multidimensional scaling) in R. Spearman rank correlation between the first axis of microbiome variation, Shannon diversity, and key functional genes and fecal calprotectin was performed using *cor*.*test* function in R.

To calculate the proportion of variance explained by each tested variable on Bray-Curtis dissimilarity (for species, pathways and enzymes), permutational multivariate analysis of variance (PERMANOVA) test was performed using *adonis* function in the R package ‘vegan’. The variables included disease phenotype, age, antibiotic use, fecal calprotectin, gender, disease activity, pouch phenotype and inter-subject variation (the last four variables were only available for the cohort of patients with a pouch and were fitted separately). The variance explained by each variable was calculated independently of other variables to avoid issues related to variable ordering.

To test for differently abundant microbiome features (species, enzymes, and pathways profiles) in different IBD phenotypes, generalized linear mixed-effects models from the R package MaAsLin2 v0.99.12 (http://huttenhower.sph.harvard.edumaaslin) were used. Bacterial taxonomy and gene relative abundance data were log-transformed and additive smoothing of minimum non-zero values was applied to zero values on a per-sample basis. The transformed abundances were modeled as a function of IBD phenotype (healthy subjects as the reference point), while adjusting for age and antibiotic use (fixed effects). For antibiotic use, a time window of 1-month (recent use) or 6-months off antibiotics was used. To account for the longitudinal dataset of pouch patients (repeated measurements), individual patients from which a set of samples were derived, were specified as a random effect (patient ID). To account for potential confounders due to the combination of several cohorts of patients, the cohort from which each sample was derived was specified as another random effect (cohort ID). All reported P values were adjusted for multiple hypotheses testing using the Benjamini-Hochberg method. For differential abundance analysis of key bacterial genes (*but, buk, atoAD, 4hbt* and *bai* genes cluster) in IBD phenotypes, a linear mixed-effects model was used with the same fixed and random effects as described above, with *lmer* function from the R package ‘lme4’.

### Machine learning classifiers

Profiles of bacterial species, metabolic pathways, and enzymes composition were used to build classification models to distinguish between pouch phenotypes (normal pouch vs. pouchitis). The machine-learning algorithm of gradient boosting trees (GBT) was used, as implemented in Python package XGBoost (eXtreme Gradient Boosting [28]) with *XGBClassifier* function. The best hyperparameters were tuned empirically using grid search matrix (*GridSearchCV* function in Python “scikit-learn” package). Briefly, learning rate = 0.025, number of boosted trees = 500 (*n_estimators*), each tree was constructed randomly with 50% of the features (*colsample_bytree*) and trained randomly on 50% of the samples (*subsample*) and a maximum depth of a tree = 3 (*max_depth*). The rest of the classifier parameters were left as in default (unchanged). The GBT classifiers were trained and evaluated on the discovery cohort (pouch samples from the normal pouch and pouchitis groups) by using five-fold cross-validation (randomly repeated 100 times) with *StratifiedKFold* function (“scikit-learn” package), that preserves the same distribution of classes in each fold as in the complete training set. The classifier was tested on the validation cohort of patients with recurrent-acute pouchitis, to test its ability to correctly predict whether these patients will become normal pouch or pouchitis in future follow-up (the true label was the phenotype defined during the last follow-up clinic visit). The model performance was assessed by calculation area under the curve (AUC), sensitivity, specificity, and accuracy metrics.

To improve the model’s predictive power (considering only the most informative features) and reduce complexity (reducing the feature space by removing uninformative and co-correlated features), feature importance scores were computed using the internal XGBoost function (feature_importances_), which assigns a score to each feature used in the decision tree during model training. The average feature importance scores for each model (species, pathways and enzymes) were calculated by considering the mean score of each feature for each fold across the repeated (x100) five-fold cross-validation.

### Availability of data and materials

All the metagenomic sequence data generated and used in this study (patients with a pouch) have been deposited in NCBI SRA and is available under BioProject number PRJNA637365. This study also used our previously published sequence data from PRJNA524170. Metagenomes from patients with UC, CD and healthy controls were obtained from PRJNA400072. Analysis scripts with code for the machine learning classifiers are available in the GitHub repository (https://github.com/VadimDu/pouchitis_classifiers).

## Results and discussion

We enrolled 75 patients with a pouch in this study; the median age was 43.8 (interquartile range 33.8-62.2), and 60% were males (Supplementary Table 1A, B). 250 fecal samples were collected longitudinally (median of 3 samples per patient) over a period of up to 5.8 years. Based on our classification system (Materials and Methods), these patients were divided into two main groups that were used throughout most analysis (discovery cohort): patients with a normal pouch (non-inflamed; n=35) and pouchitis (inflamed; n=34). The normal pouch and pouchitis groups consisted of 103 and 105 samples, respectively. An additional 42 samples were obtained from patients with recurrent-acute pouchitis and were used as a validation cohort in order to test our machine-learning classifier predictions of pouchitis occurrence based on microbiome data. Patients with recurrent-acute pouchitis suffer from up to 4 flares of pouchitis per year.

To explore the microbiomes of patients with a pouch in the context of primary IBD, we analyzed shotgun metagenomes derived from fecal samples of patients with a pouch, UC, CD and healthy subjects in terms of species composition and functional gene repertoires (Materials and Methods). Metagenomes of patients with CD (n=88), UC (n=76) and of healthy subjects (n=56) were obtained from data previously published in [18], which are part of the PRISM, LifeLines DEEP and NLIBD cohorts (cross-sectional) (Supplementary Table 1C). To minimize potential biases, raw metagenomic reads from all cohorts were processed in parallel through the same bioinformatic pipeline, in line with recent recommendations for multi-cohort studies (^29^Szamosi et al. 2020).

### Microbial dysbiosis across disease phenotypes and its association with inflammation

The microbial community composition among disease phenotypes and healthy subjects (beta-diversity, analyzed by PCoA) was well separated according to profiles of species, genes and pathways (Fig. 1A-C), with some degree of overlap, in particular between samples from UC and CD patients as was observed previously [18]. Strikingly, we observed a significant stratification (Kruskal-Wallis P < 0.01) of different disease phenotypes based on the first axis of microbiome variation, shifting away from the healthy subjects, in the following order: UC, CD, normal pouch, and finally patients with pouchitis (Fig. 1A-C). The latter was the most distant from the microbiomes of healthy controls, i.e. with the highest level of dysbiosis. Interestingly, according to Shannon diversity (Fig. 1D), no significant difference was observed between patients with CD (median Shannon 2.6) and UC (median Shannon 2.87) to patients with a normal pouch (median Shannon 2.73). In contrast, patients with pouchitis had significantly lower diversity (median Shannon 2.31, Kruskal-Wallis P < 0.001) than all other phenotypes. Patients with UC had significantly higher diversity than those with CD (Kruskal-Wallis P = 0.0044) and only slightly lower diversity than that of healthy subjects (median Shannon 2.96, Kruskal-Wallis P = 0.004), in agreement with previous reports [7].

**Fig. 1.**
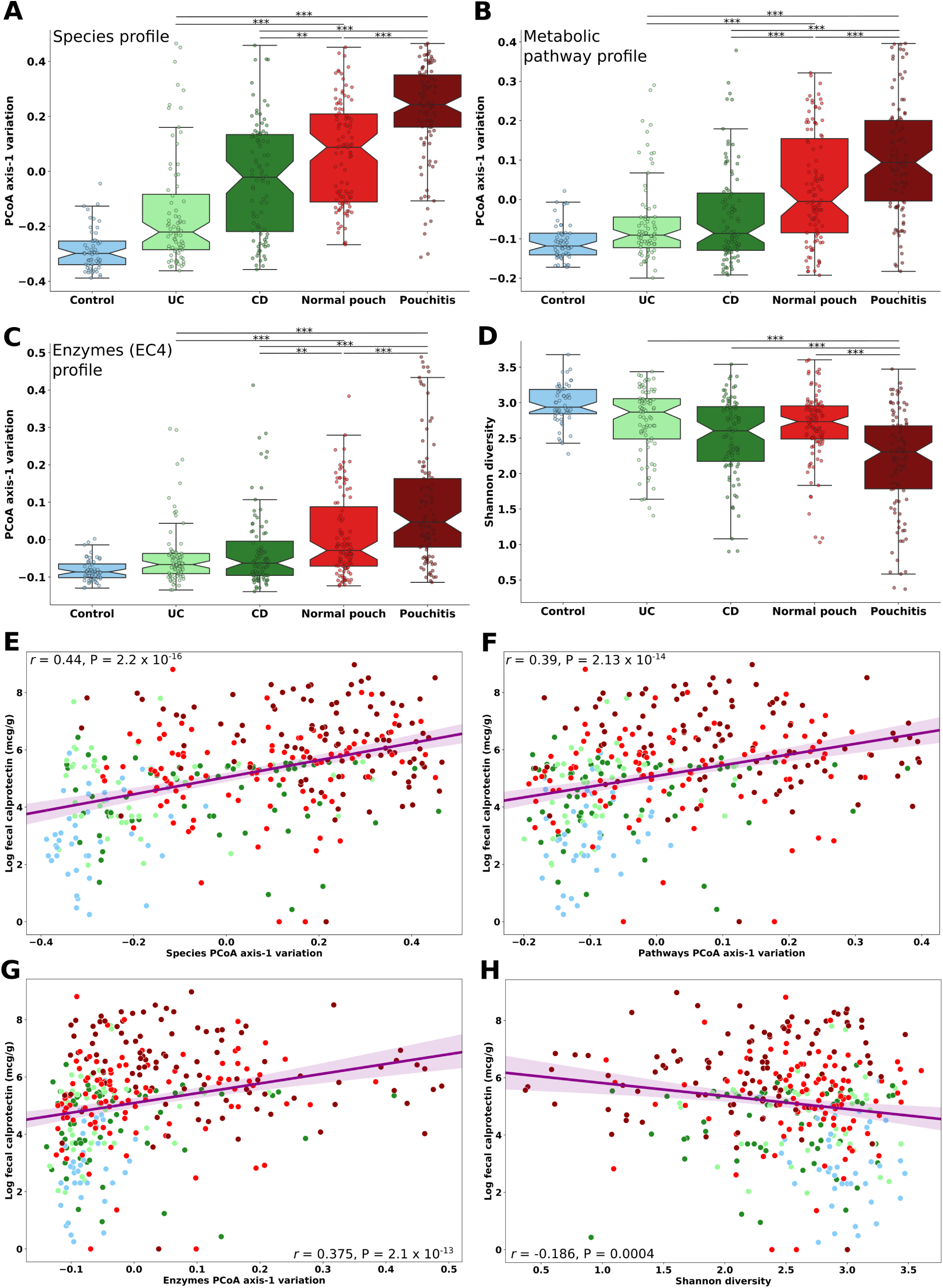
Microbial community stratification among disease phenotypes and healthy subjects. The microbiome variation according to the first axes of principal coordinates analysis (PCoA) of (A) species, (B) metabolic pathways and (C) enzymes profiles, based on Bray-Curtis dissimilarity of the fecal metagenomes. (D) Shannon diversity in the gut microbiome based on species composition. (E - H) Spearman correlation calculated between fecal calprotectin level (log) and the first axes of microbiome variation for species (E), metabolic pathways (F), enzymes (G) and Shannon diversity (H). Colors represent the different IBD phenotypes (UC, CD, Normal pouch and Pouchitis), as well as healthy subjects (Control) study groups. *P < 0.05; **P < 0.01; ***P < 0.001; Kruskal-Wallis, applying Dunn’s multiple correction test. For the list of all pairwise P-values see Supplementary Table 2. Boxplot whiskers mark observations within the 1.5 interquartile range of the upper and lower quartiles.

The subject’s clinical phenotype explained a modest part of the observed variation in microbial composition (beta-diversity based on Bray-Curtis distance) in species, metabolic pathway and enzymes composition (Supplementary Fig. 1; PERMANOVA, R^2^ = 12.1%, 13.2%, 10.4%4 respectively; P < 0.001), while patient age and gender (available only for the pouch cohort) explained only 0.39% to 0.71% and 0.42% to 0.52%, respectively. Antibiotic use (1-month or 6-months off-antibiotics) accounted for only 0.45% to 0.62% of the variation. Fecal calprotectin, a biomarker for intestinal inflammation, explained a low yet significant degree of microbial variation (R^2^ = 2.5% to 4.1%, P < 0.001). Consistent with previous studies of patients with UC and CD [30, 13], inter-subject variation explained the highest degree of microbiome variation (Supplementary Fig. 1D; PERMANOVA, R^2^ = 46.6% to 48.7%, P < 0.001) among the longitudinal samples of patients with a pouch cohort. Finally, in patients with a pouch cohort, pouch phenotype and disease activity contributed 4.2% to 4.8% and 1.4% to 1.9%, respectively, to the variation in microbial species, pathways and enzymes (Supplementary Fig. 1D). These observations indicate that besides inter-subject differences, disease type and inflammation severity are the strongest contributors to microbial variability and the resulting dysbiosis.

If dysbiosis is associated with intestinal inflammation, one would expect a correlation between the microbiome variation described above and inflammation biomarkers. Indeed, we observed a moderate and significant correlation between the fecal calprotectin level and the first axis of microbiome variation for species, metabolic pathways and enzymes (Fig. 1E-G; Spearman *r* = 0.44, 0.39 and 0.375 respectively; P < 0.001). A weaker negative correlation was detected between calprotectin and Shannon diversity (Fig. 1H; Spearman *r* = −0.186, P = 4.01 x 10^−4^). Patients with pouchitis had the highest level of calprotectin, followed by patients with a normal pouch. Patients with CD and UC had similar median calprotectin levels (Supplementary Fig. 2). Altogether these data suggest that patients with a normal pouch already show signs of inflammation and dysbiosis that may parallel those observed in CD, and that these are intensified in patients with pouchitis.

To rule out the possibility that the observed differences between the pouch to non-pouch patients microbiomes are inflated due to center or country effects, we obtained 15 publicly available pouch metagenomes [31] (an American cohort) and compared alpha and beta diversity across all the analyzed metagenomes. Reassuringly, based on both microbiome species variation (Bray-Curtis) and Shannon diversity (Supplementary Fig. 3), the pouch samples from [31] clustered within the range of the pouches from our RMC pouch cohort.

### Bacterial species associated with IBD

We aimed to characterize the altered species composition in the four IBD groups compared to healthy subjects, and also to compare patients with a pouch to primary IBD, using linear modeling of shotgun metagenomic data. As expected, bacterial species composition showed extensive and significant differences involving many taxa between healthy subjects and patients with UC, CD and with a pouch (Fig. 2, Supplementary Table 3). We confirmed past observations of decreased abundance in beneficial taxa such as *Faecalibacterium prausnitzii, Eubacterium rectale* and several *Roseburia* and *Bacteroides* species and increased abundance of potential pathobionts such as *Escherichia coli, Pantoea* sp., and several *Veillonella* and *Streptococcus* species in CD and in patients with a pouch [8, 10, 11, 12]. Patients with a pouch (in particular those with pouchitis) showed the strongest extremes in terms of the levels of the above mentioned genera, further supporting the existence of a strong dysbiosis in pouchitis.

**Fig. 2.**
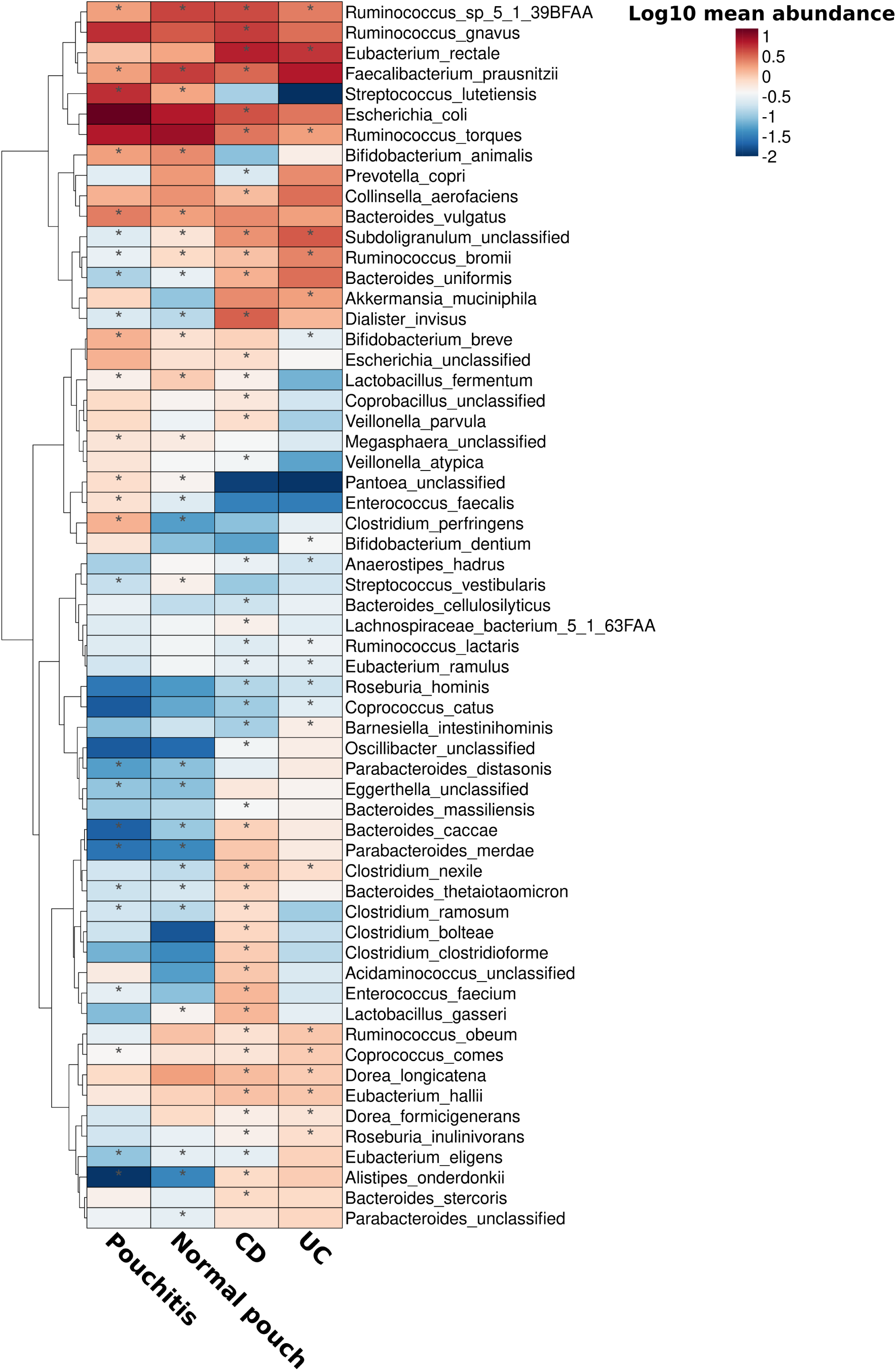
Bacterial species associated with IBD phenotypes. A generalized linear mixed-effects model was built for each bacterial species, and IBD phenotypes were set as a predictor (including age and antibiotic use as fixed effects). Healthy subjects were used as a reference point, and individual subjects and cohort were set as random effects. Only species with a mean relative abundance across all groups ≥ 0.15% with a least one significant association with an IBD phenotype are presented. For the full list of species that were significantly associated with IBD phenotypes see Supplementary Table 2. Asterisks indicate patient groups for which the association was significant (FDR < 0.05).

In addition, we identified significant depletion of *Subdoligranulum* in all disease phenotypes (Fig. 2), with the lowest abundance in patients with a pouch (normal pouch FDR P = 0.025, pouchitis FDR P = 0.01). Species belonging to this genus, which is closely related to *Faecalibacterium*, have been shown to produce the anti-inflammatory SCFA butyrate [32] and consequently may play a protective role in the gut. Additionally, *Ruminococcus bromii*, a keystone species that is highly efficient in the degradation of resistant starch, making its degradation products available for other beneficial bacteria via cross-feeding [33], was highly depleted in all disease phenotypes, especially in patients with a pouch (normal pouch FDR P = 5.7 x 10^−7^, pouchitis FDR P = 7.5 x 10^−9^). Interestingly, we found that *Ruminococcus gnavus* and *Ruminococcus torques* were enriched in patients with a pouch (not statistically significant), while *Akkermansia muciniphila* was depleted (Fig. 2, normal pouch FDR P = 0.12, pouchitis FDR P = 0.15). These three species have the potential to degrade human secretory mucin, and a similar trend of increase in *R. gnavus* and *R. torques* coupled to a decrease in *A. muciniphila* was previously reported in biopsies of UC and CD patients [34]. A higher proportion of *R. gnavus* and *R. torques* in patients with a pouch may be an indication of excessive mucin degradation in the pouch, which may contribute to increased intestinal inflammation [35]. Notably, a distinct clade of *R. gnavus* strains was previously found to be enriched in CD and UC patients (marked by considerable yet transient blooms) and harbored potential virulence-associated genes [36]. To critically examine the hypothesis of increased mucin degradation potential in the pouch, we moved on to quantify the total mucin-degrading bacterial enzymes in the gut microbiomes of these patients.

### Mucin degradation and R. gnavus glycoside hydrolases

We specifically analyzed glycoside hydrolase genes (GH) that are involved in the breakdown and utilization of mucin-derived glycans and are encoded in the genomes of mucolytic bacteria. According to these criteria, bacterial community mucin degradation potential in patients with a normal pouch was similar to that of patients with CD (Kruskal-Wallis, P = 0.08) and to healthy subjects (Fig. 3A; Kruskal-Wallis, P = 0.374). Patients with pouchitis had 12.3% lower community mucin degradation potential than patients with a normal pouch (Kruskal-Wallis, P = 3.4 x 10^−4^), while patients with UC had the lowest mucin degradation potential than the other phenotypes (Fig. 3A; Kruskal-Wallis, P < 0.01).

**Fig. 3.**
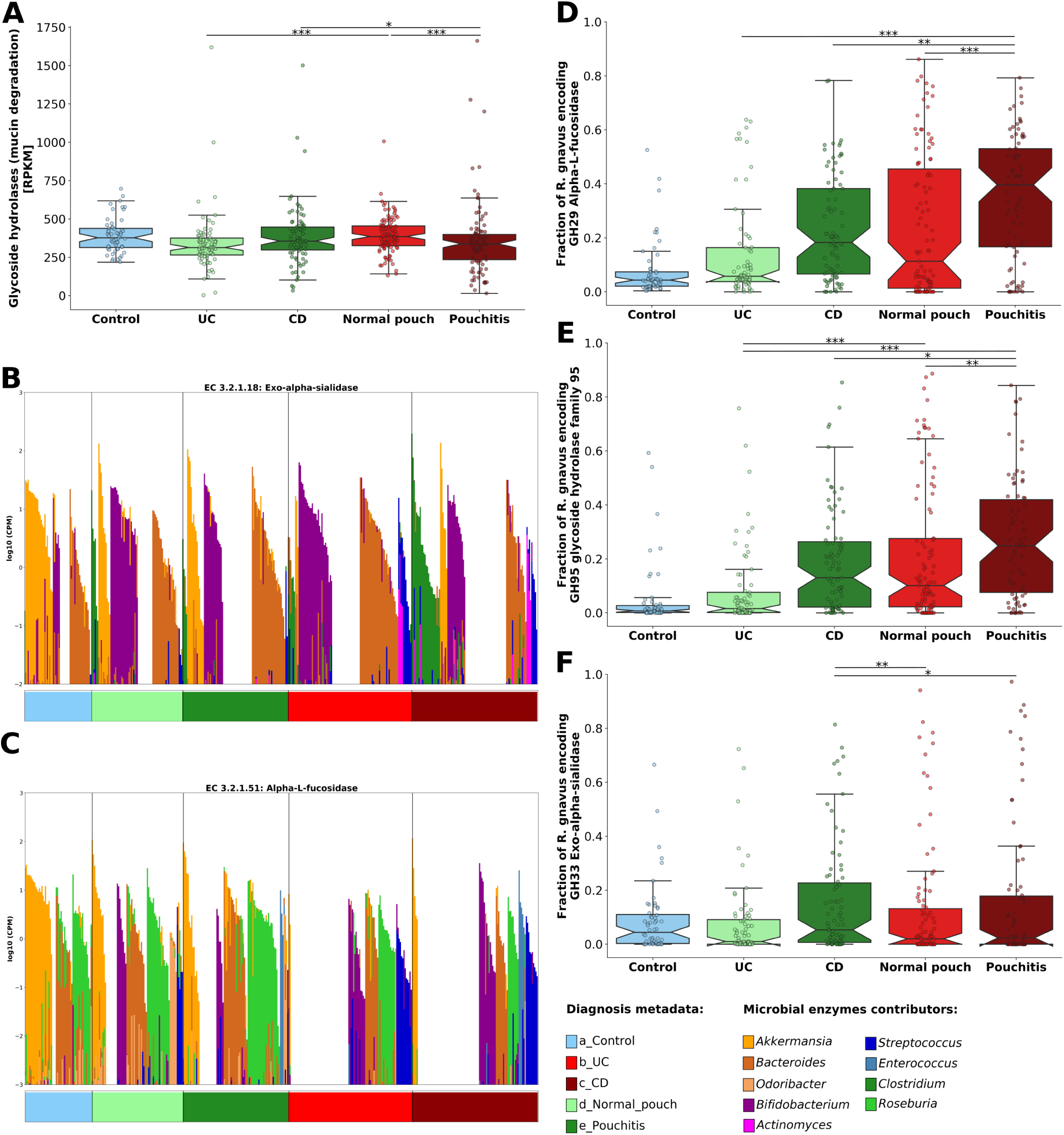
Mucin degradation potential of the microbiome of patients with IBD and healthy controls. (**A**) Bacterial community mucin degradation potential based on analysis of glycoside hydrolase genes (GH) that are involved in the breakdown and utilization of mucin-derived glycans. (**B, C**) Contribution of glycoside hydrolase enzymes Alpha-L-fucosidase and Exo-alpha-sialidase by bacterial genera. Each bar on the x-axis represents a sample from the corresponding phenotype (color coded) and the y-axis is the log-scaled total community gene abundance (sum-normalized to copies per million [cpm] units). The contributions of the top genera are proportionally scaled within the total, see the legend at the bottom of the figure (**D** - **F**) The relative fraction of *Ruminococcus gnavus* strains encoding strain-specific glycoside hydrolases (normalized according to three single-copy housekeeping genes) involved in mucin degradation; GH29 Alpha-L-fucosidase (**D**), GH95 glycoside hydrolase family 95 (**E**) and GH33 Exo-alpha-sialidase (**F**). *P < 0.05; **P < 0.01; ***P < 0.001; Kruskal-Wallis, applying Dunn’s multiple correction test. For the list of all pairwise P-values see Supplementary Table 2. Boxplot whiskers mark observations within the 1.5 interquartile range of the upper and lower quartiles.

To better understand the specific role of *R. gnavus* in mucin degradation, we focused on its enzymes that are considered specific for host mucin utilization: GH29 (Alpha-L-fucosidase), GH95 (glycoside hydrolase family 95) and GH33 (Exo-alpha-sialidase). The former two can cleave fucose from host mucin glycans while the latter can cleave terminal sialic acid from sialylated mucins. It was shown that the ability of *R. gnavus* to utilize host mucins is strain-dependent and is attributed to a specific GH33 sialidase, and to GH29 and GH95 fucosidases [^37^Crost et al. 2013]. Current HUMAnN2 enzyme annotation lacks *R. gnavus* as a contributor to Alpha-L-fucosidase (EC 3.2.1.51) and Exo-alpha-sialidase (EC 3.2.1.18) enzymes (Fig. 3B,C). Thus we obtained sequences of GH29, GH95 and GH33 from the representative genome of *Ruminococcus gnavus* ATCC-29149 JCM6515 and quantified the abundance of these genes and the fraction of *R. gnavus* species that possess them in the metagenomes by read-mapping. The fraction of *R. gnavus* strains encoding GH29 and GH95 fucosidases varied extensively across disease phenotypes (Fig. 3D,E), but was significantly higher in patients with pouchitis with 39.6% and 25% (median) of the strains encoding these fucosidases, respectively (Kruskal-Wallis, P < 0.01). No significant difference was observed between patients with a normal pouch (11.3% and 10.1% median) and CD (18.3% and 13% median) for GH29 and GH95, respectively. Less than 6% of *R. gnavus* in UC patients and in healthy subjects encoded these fucosidases (Fig. 3D,E). Interestingly, GH33 sialidase was encoded by a significantly smaller proportion of *R. gnavus* strains (Fig. 3F). Patients with a pouch had a similar fraction of *R. gnavu*s-encoded GH33 sialidase (normal pouch 2% and pouchitis 2.4% medians; Kruskal-Wallis, P = 0.336), which was comparable to UC patients and healthy subjects (Kruskal-Wallis, P > 0.05). Patients with CD had the highest abundance of *R. gnavus* strains encoding GH33 sialidase (Fig. 3F; 5.3% median; Kruskal-Wallis, P < 0.05).

A study by [38] established that the pH level in patients with a normal pouch (5.4) was significantly lower than in patients with pouchitis (6.5). Remarkably, this low pH in the normal pouch substantially inhibited mucin (including fucose residues) degradation. This might explain why we detected a markedly lower proportion of *R. gnavus* strains encoding fucosidases in patients with a normal pouch. Altogether, our findings demonstrate that as a consequence of strong dysbiosis in pouchitis, the overall bacterial community mucin degradation potential is lower, but there is a significantly higher number of mucin-degrading *R. gnavus* strains that potentially might be pro-inflammatory. Mucin degradation in a normal pouch is similar to that of CD both in terms of microbial community- and *R. gnavus*-associated potential. These findings highlight the importance of mucin degradation in intestinal inflammation, as well as the possible effect of disease phenotype and location (whether small or large intestine).

### Butyrate synthesis potential of the microbiome is deprived in pouchitis

Following our finding that patients with a pouch had greatly reduced abundances of beneficial species, some of which are known SCFA producers, we aimed to directly quantify the genes encoding SCFA synthesis enzymes. SCFA are the major metabolic products of anaerobic fermentation of non-digestible polysaccharides by the human colonic microbiota [39, 40]. Here we focused on butyrate, as it is considered to have the greatest benefit to the human host. Butyrate is the preferred energy source for colonocytes, improves the gut barrier function, has immunomodulatory and anti-inflammatory properties and reduces oxidative stress in the colon [39]. Butyrate-producing species are found among many butyrate non-producing species in two dominant families of Firmicutes, *Ruminococcaceae* and *Lachnospiraceae* [40]. Four main pathways are known for butyrate production: the acetyl-CoA, glutarate, 4-aminobutyrate, and lysine pathways. Acetyl-CoA is the dominant pathway for butyrate synthesis and is present in the majority of butyrate producers and *but* is the most prevalent terminal gene [41].

We quantified and modeled the terminal genes of the four known butyrate production pathways, namely *but, buk, ato* and *4hbt* (see Materials and Methods) in the metagenomes using an extensive and rigorously-curated database for butyrate synthesis genes [24]. Overall, patients with CD, normal pouch and pouchitis had significantly lower levels of butyrate synthesis pathways compared to healthy subjects (Fig. 4A-D; P < 0.05). Specifically, according to the *but* gene (Fig. 4A, E), the lowest abundance of these genes was in patients with pouchitis (median RPKM 12), followed by significantly higher abundance in patients with a normal pouch (median RPKM 78; Kruskal-Wallis, P = 3.5 x 10^−6^). No significant difference in butyrate synthesis genes between CD (median RPKM 126) and UC (median RPKM 136) was observed, but both were significantly lower than in healthy subjects (median RPKM 175; Kruskal-Wallis, P = 3 x 10^−5^, P = 0.002, respectively) and higher than in pouch phenotypes. Analysis of the *buk* homologs (Fig. 4B, F) revealed no significant difference in abundance between patients with CD (median RPKM 6.6) and with normal pouch (median RPKM 8.4) or with pouchitis (median RPKM 4.75), but patients with pouchitis had a lower abundance of *buk* than with normal pouch (Kruskal-Wallis, P = 0.025). Patients with UC and healthy subjects had the highest gene abundance (Kruskal-Wallis, P < 0.01). The lysine (*ato*) and 4-aminobutyrate (*4hbt*) pathways were negatively associated with patients with a pouch (Fig. 4C, D), and were less abundant in general in all groups, presenting similar trends (Fig. 4G, H). Patients with a pouch had very low levels of both *ato* and *4hbt* genes (were missing altogether in many samples) suggesting that the 4-aminobutyrate and lysine pathways for butyrate synthesis are exceptionally low in these patients.

**Fig. 4.**
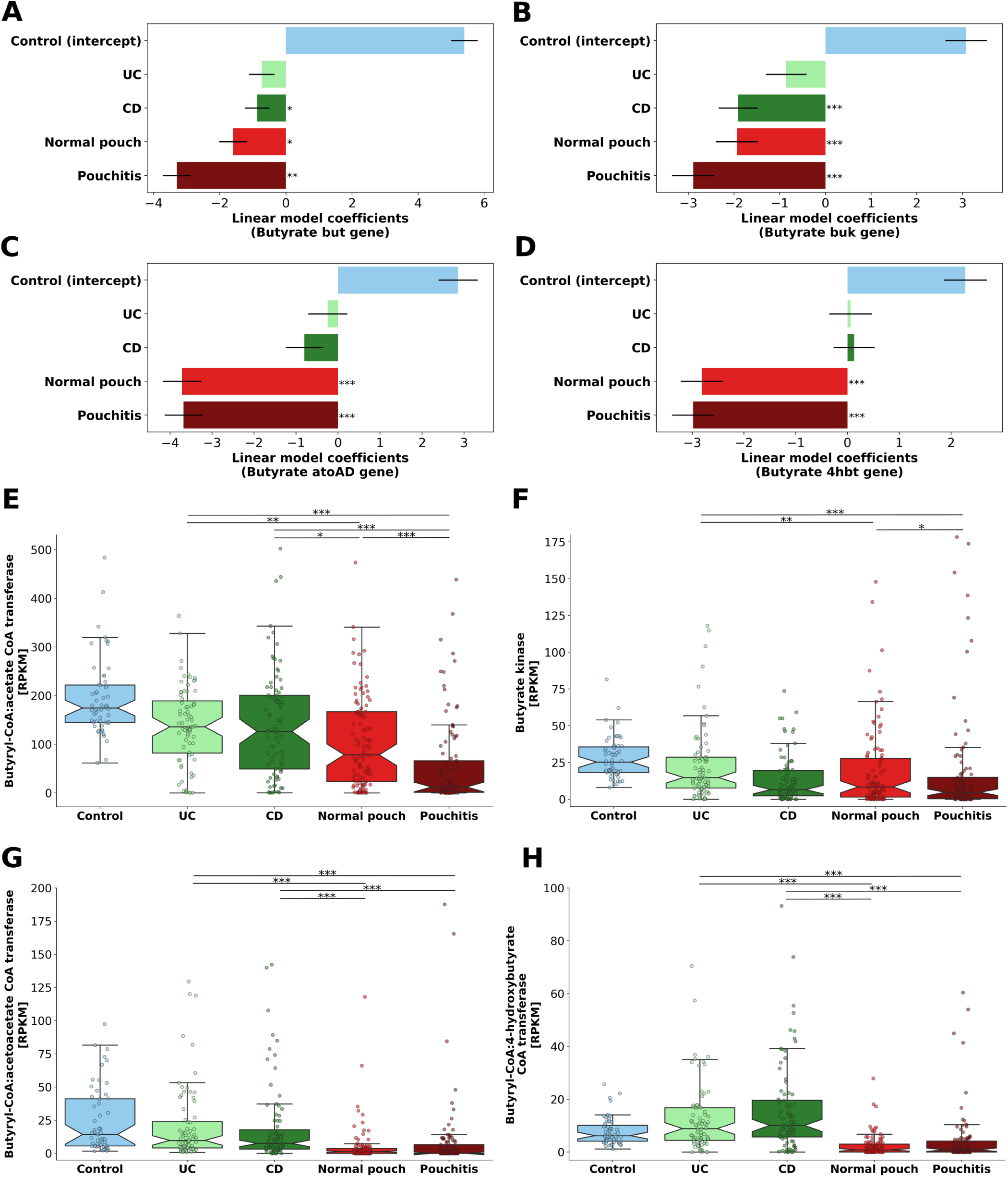
The main pathways of microbial butyrate synthesis in IBD and healthy subjects. Modeling and quantification of the terminal genes (*but, buk, ato* and *4hbt*) for corresponding butyrate synthesis pathways in the metagenomes: (**A, E**) butyryl-CoA:acetate CoA transferase (*but*); (**B, F**) butyrate kinase (*buk*) - acetyl-CoA and glutarate pathways; (**C, G**) butyryl-CoA:acetoacetate CoA transferase (*ato*) -lysine pathway; (**D, H**) butyryl-CoA:4-hydroxybutyrate CoA transferase (*4hbt*) - 4-aminobutyrate pathway. In **A**-**D**, linear model coefficients (effect sizes) are given for each phenotype for the modeled genes. *P < 0.05; **P < 0.01; ***P < 0.001. In **E**-**H**, gene abundance is in RPKM units (reads per kilobase per million mapped reads), normalized according to the corresponding median gene length and total-sum scaled to the number of reads in each metagenome. *P < 0.05; **P < 0.01; ***P < 0.001; Kruskal-Wallis, applying Dunn’s multiple correction test. For the list of all pairwise P-values see Supplementary Table 2. Boxplot whiskers mark observations within the 1.5 interquartile range of the upper and lower quartiles.

The taxonomic affiliation of butyrate-producing bacteria based on the similarity of the identified *but* and *buk* genes to known butyrate producers revealed a dominance of commensal Firmicutes such as *F. prausnitzii* and several species of *Roseburia* and *Eubacterium* (Supplementary Fig. 4). Surprisingly, healthy subjects had a somewhat low relative abundance of some of these butyrate producers compared to disease phenotypes, which might be a result of the higher bacterial diversity in healthy, and consequently a lower “reliance” on a few species for butyrate production. *F. prausnitzii, E. halii*, and *Subdoligranulum variabile* had comparable relative abundances in patients with normal pouch, UC and healthy, while patients with pouchitis and CD had low levels of these taxa. For a complete list of all the identified butyrate producers, see Supplementary Table 4. Interestingly, *ato*-encoding *Alistipes putredinis* and *Odoribacter splanchnicus* (Bacteroidetes), were abundant only in non-pouch patients. Conversely, *Cetobacterium somerae* (Fusobacteria) encoding *ato* was highly abundant in seven samples from patients with a pouch and in only a single sample from a CD patient. Notably, a significant negative correlation was observed between the abundance of butyrate synthesis genes and fecal calprotectin (Supplementary Fig. 5). This was most pronounced for the *but* gene (Spearman *r* = −0.322, P = 4.34 x 10^−10^). This suggests that either a reduction in butyrate producers predisposes to intestinal inflammation or that butyrate-producing bacteria are more sensitive to intestinal inflammation.

As butyrate production may be affected by patients’ dietary habits, especially dietary fiber consumption, we correlated the average daily intake of dietary fiber derived from different food sources with butyrate synthesis genes in a subset of samples (n=52) that had dietary information (FFQs obtained at the time of fecal samples collection) - Table 1. The abundance of *but* and *buk* genes were positively correlated with total dietary fiber intake (Spearman *r* = 0.366, P = 0.0076), and specifically with intake of fiber from fruit (Spearman *r* = 0.293, P = 0.035), vegetables (Spearman *r* = 0.275, P = 0.048) and potatoes (Spearman *r* = 0.46, P = 6 x 10^−4^). Higher fruit consumption had been previously associated with increased abundance of known butyrate-producing species, and lower recurrence of pouchitis [42]. Nutritional intervention for patients with a pouch, such as the Mediterranean diet was previously associated with decreased inflammatory markers and later onset of pouchitis and is characterized by a high intake of dietary fibers [43]. Our results, therefore highlight diet as an important factor that may affect butyrate synthesis and provide further support for dietary intervention as a plausible approach to modulate the microbiota and increase the low butyrate levels in the pouch.

Previous studies examining butyrate-producing bacteria, their gene content or metabolites in IBD reported various results. A study of [44] reported no difference in butyrate concentrations in fecal samples between healthy subjects and UC patients. On the other hand, fecal butyrate concentrations were decreased in both CD and (to a lesser degree) UC patients in earlier studies [45, 46]. According to the quantification of *but* gene content, reduced butyrate synthesis was identified in patients with active and inactive CD but only in patients with active UC [47]. In patients with a pouch, data about butyrate production is scarce and butyrate synthesis genes had not been previously analyzed. It was suggested that bacterial dysbiosis in pouchitis causes SCFA deprivation [6], which might be related to the pathogenesis of pouchitis. Fecal levels of SCFA in patients with pouchitis were substantially lower compared to patients with a normal pouch [48, 49], which may contribute to the higher pH observed in pouchitis [38]. These past observations of butyrate and pH levels in the pouch are in line with our metagenomics-based findings and suggest that a shortage of butyrate (and its producers) is an important contributor to the pathogenesis of pouchitis. This may be clinically relevant as SCFA enemas were successfully assessed for the treatment of diversion colitis [50] and for UC [51]. Their effect in pouchitis was rarely assessed in small uncontrolled trials and case studies [52]. Our findings provide a mechanistic rationale for a reassessment of this therapy in pouchitis.

### Secondary bile acids production potential is low in pouchitis

Secondary bile acids deoxycholic acid (DCA) and lithocholic acid (LCA) are formed in the colon from primary bile acids metabolism by the microbiota via a multi-step 7α-dehydroxylation reaction. Secondary bile acids (DCA and LCA) have extensive effects on host metabolism and play both negative and positive roles in health and disease [53]. The level of DCA in bile is thought to be controlled by two major factors: levels and activities of bile acid 7α-dehydroxylating gut microbes and colonic transit time [53]. In IBD, impaired fecal bile acid metabolism was observed, with an increase in primary bile acids and a decrease in secondary bile acids (particularly during flares). *In-vitro* and *in-vivo* experimental models confirmed that DCA and LCA may exert anti-inflammatory effects [54, 55]. Enzymes involved in 7α-dehydroxylation are encoded by bile acid inducible (*bai)* gene clusters present in bacterial genomes from *Ruminococcaceae* and to a lesser extent from *Lachnospiraceae* and *Peptostreptococcaceae* [56]. The *bai* genes cluster was found to be encoded and expressed in the gut microbiomes of most individuals, however only in a small fraction (<1%) of total intestinal bacteria [25].

We quantified and modeled the *bai* genes cluster (see Materials and Methods) in the metagenomes of IBD and healthy subjects using an extensive and rigorously curated database for secondary bile acids formation genes [25]. Patients with CD, normal pouch, and pouchitis were significantly associated with a decreased secondary bile acid formation potential (Fig. 5A; P < 0.05). In line with the highest dysbiosis in patients with pouchitis, the *bai* gene cluster abundance was lowest in this group (median RPKM 21.3), but was higher in patients with a normal pouch (Fig. 5B; median RPKM 44.6; Kruskal-Wallis, P = 0.002). UC and CD patients had comparable levels of *bai* genes abundance (median RPKM 72.6 and 71, respectively; Kruskal-Wallis, P = 0.29), and both were significantly higher than in patients with pouch (Kruskal-Wallis, P < 0.001). Healthy individuals had the highest abundance of *bai* genes of all phenotypes (Fig. 5B; median RPKM 101; Kruskal-Wallis, P < 0.01).

**Fig. 5.**
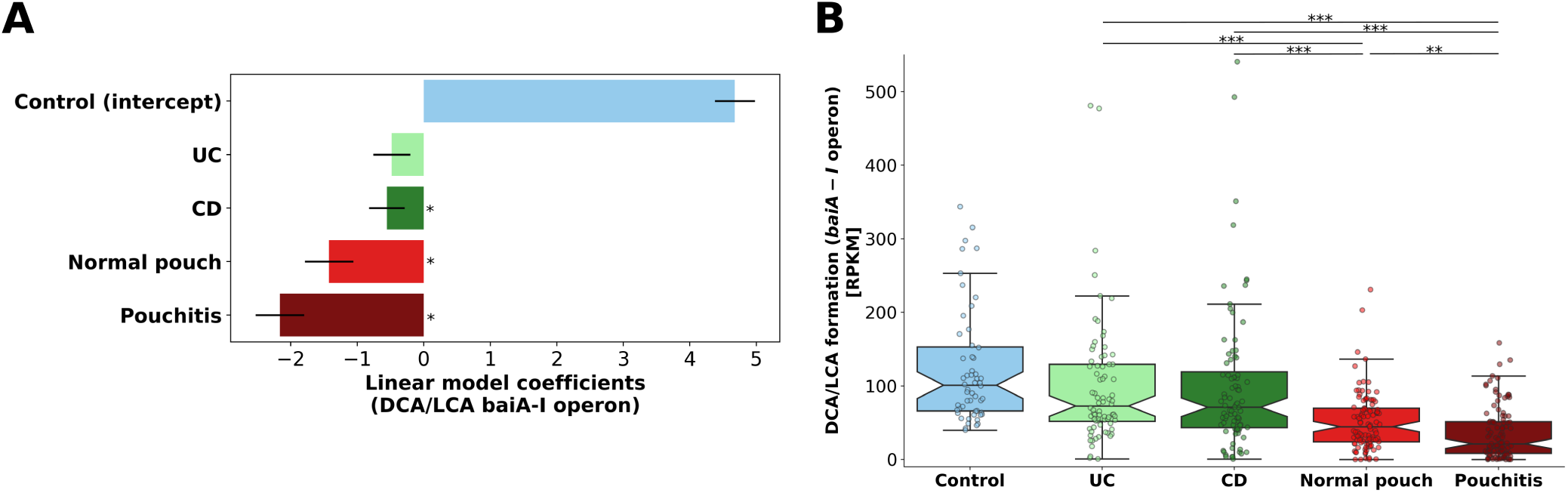
Secondary bile acids (DCA and LCA) formation potential in the metagenomes of patients with IBD and healthy subjects. The *bai* genes cluster (*baiA-I* operon) was quantified in the metagenomes. (**A**) Linear model coefficients (effect sizes) are given for each phenotype for the modeled *bai* genes cluster. *P < 0.05 (**B**) Gene abundance is in RPKM units (see legend of Fig. 4). *P < 0.05, **P < 0.01; *P < 0.01; Kruskal-Wallis, applying Dunn’s multiple correction test. For the list of all pairwise P-values see Supplementary Table 2. Boxplot whiskers mark observations within the 1.5 interquartile range of the upper and lower quartiles.

Only several dominant strains in the analyzed metagenomes were identified to encode the *bai* gene clusters (or parts of). They belonged to three families of Firmicutes: *Lachnospiraceae* (*Clostridium scindens*), *Peptostreptococcaceae* (*Clostridium sordellii* and *Proteocatella sphenisci*) and *Ruminococcaceae* (uncultured strains of Firmicutes bacterium CAG:103, Clostridiales bacterium UBA4701 and UBA1701). For a complete list of the identified secondary bile acid producers, see Supplementary Table 4. Importantly, the abundance of *bai* gene cluster was significantly negatively correlated with fecal calprotectin (Supplementary Fig. 6; Spearman *r* = −0.312, P = 1.64 x 10^−9^), implying that DCA and LCA producing bacteria are associated with lower levels of intestinal inflammation.

A reduction in secondary bile acids genes in patients with CD or UC [25, 57] as well as a reduction in DCA and LCA metabolites in highly-dysbiotic patients with UC and CD, was previously described [13], supporting our current study. The key species encoding *bai* clusters in our metagenomes are concordant with those identified in metagenomes of a healthy population [56]. Importantly, one of the prevalent secondary bile acids producers we detected, *C. scindens*, was associated with enhanced resistance to *C. difficile* infection in a secondary bile acid dependent manner [58]. The reduction of secondary bile acid-producing bacteria in IBD and particularly in pouchitis might leave these patients more exposed to pathogens or potential pathobionts. Interestingly, patients with an ileal pouch were previously shown to have markedly lower levels of biliary and fecal DCA and LCA levels (compared to healthy controls), while fecal primary bile acids excretion was significantly higher, indicating attenuated bacterial conversion [59]. These measurements complement our metagenomic findings and support our hypothesis that a reduction in 7α-dehydroxylation bacterial enzymes leads to a pouch microbiome with lower potential for producing secondary bile acids.

### Enzymes related to protection against oxidative stress are highly prevalent in patients with a pouch

Previous studies suggested higher oxidative stress in the inflamed gut of patients with UC and CD [60, 9, 36]. Here we aimed to characterize and quantify genes that encode enzymes involved in oxidative stress response (oxygen-detoxifying enzymes) as a proxy for the levels of oxygen-tolerance of microbes in the metagenomes of patients with IBD. We used the enzyme profiles annotated with HUMAnN2 and extracted such oxygen-detoxifying enzymes.

We detected a significant increase in the abundance of genes encoding two enzymes responsible for the synthesis of glutathione, an important antioxidant [61], namely glutamate-cysteine ligase (EC 6.3.2.2) and glutathione synthase (EC 6.3.2.3), in patients with CD and more so in patients with a pouch (Fig 6A, B, Fig. 7A, Supplementary Table 5), again supporting similarities between these conditions. Furthermore, the gene encoding for glutathione-disulfide reductase (EC 1.8.1.7), which serves to maintain glutathione in the reduced form, was highly enriched in patients with a pouch (Fig 6C, Fig. 7A, Supplementary Table 5) (normal pouch FDR P = 9.8 x 10^−11^, pouchitis FDR P = 1.2 x 10^−13^). A trend of higher abundance of genes encoding for superoxide dismutase (EC 1.15.1.1; normal pouch FDR P = 0.65, pouchitis FDR P = 0.23, not statistically significant) and peroxiredoxin (EC 1.11.1.15; normal pouch FDR P = 0.15, pouchitis FDR P = 0.15) was detected in patients with a pouch, further supporting the notion of a pouch environment with a high oxidative stress (Fig 6D, E, Fig. 7A, Supplementary Table 5). Both of these enzyme families are known to play a central role in oxidative stress response [62, 63]. Finally, genes encoding methionine sulfoxide reductase (EC 1.8.4.11), an enzyme that reduces the oxidized form of methionine back to the active amino acid thereby reactivating damaged peptides [64], were elevated in patients with pouchitis (FDR P = 0.22, not statistically significant; Fig 6F, Fig. 7A, Supplementary Table 5). The combined contribution of these six oxidative stress response enzymes (“total oxidative stress response”) was the highest in patients with pouchitis (median RPKM 385; Kruskal-Wallis, P < 0.05), followed by patients with a normal pouch (median RPKM 282; Kruskal-Wallis, P < 0.05), and was significantly lower in the other phenotypes (Fig. 7B).

**Fig. 6.**
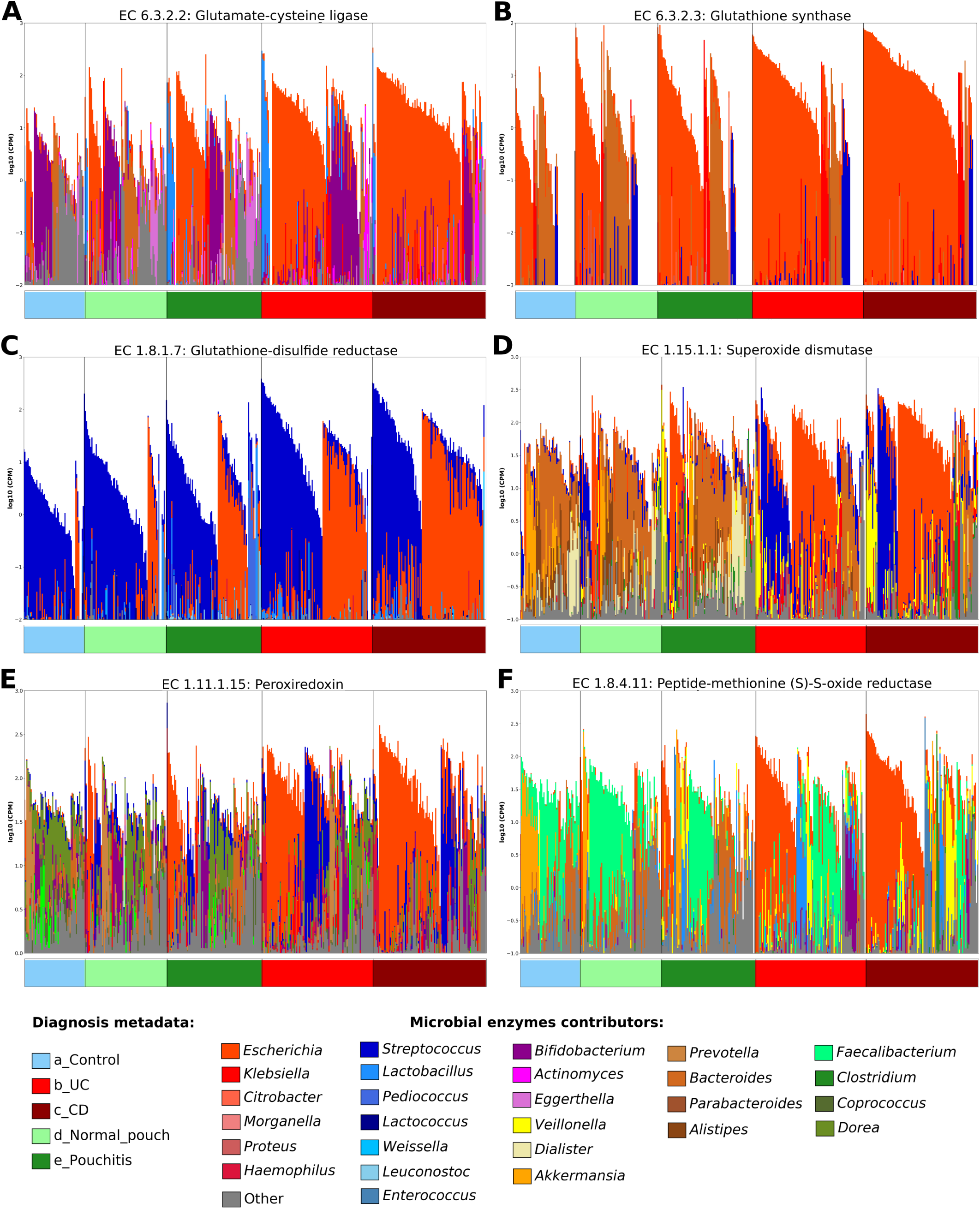
Enzymes related to protection against oxidative stress and the bacteria encoding them in the metagenomes of patients with IBD and healthy subjects. Six such enzymes were identified and quantified; (**A**) glutamate-cysteine ligase, (**B**) glutathione synthase, (**C**) glutathione-disulfide reductase, (**D**) superoxide dismutase, (**E**) peroxiredoxin and (**F**) peptide-methionine sulfoxide reductase. Each bar on the x-axis represents a sample from the corresponding phenotype (color coded) and the y-axis is log-scaled total community gene abundance (sum-normalized to copies per million [cpm] units). The contributions of the top genera are proportionally scaled within the total, “Other” encompasses contributions from additional lower abundance genera, see the legend at the bottom of the figure.

**Fig. 7.**
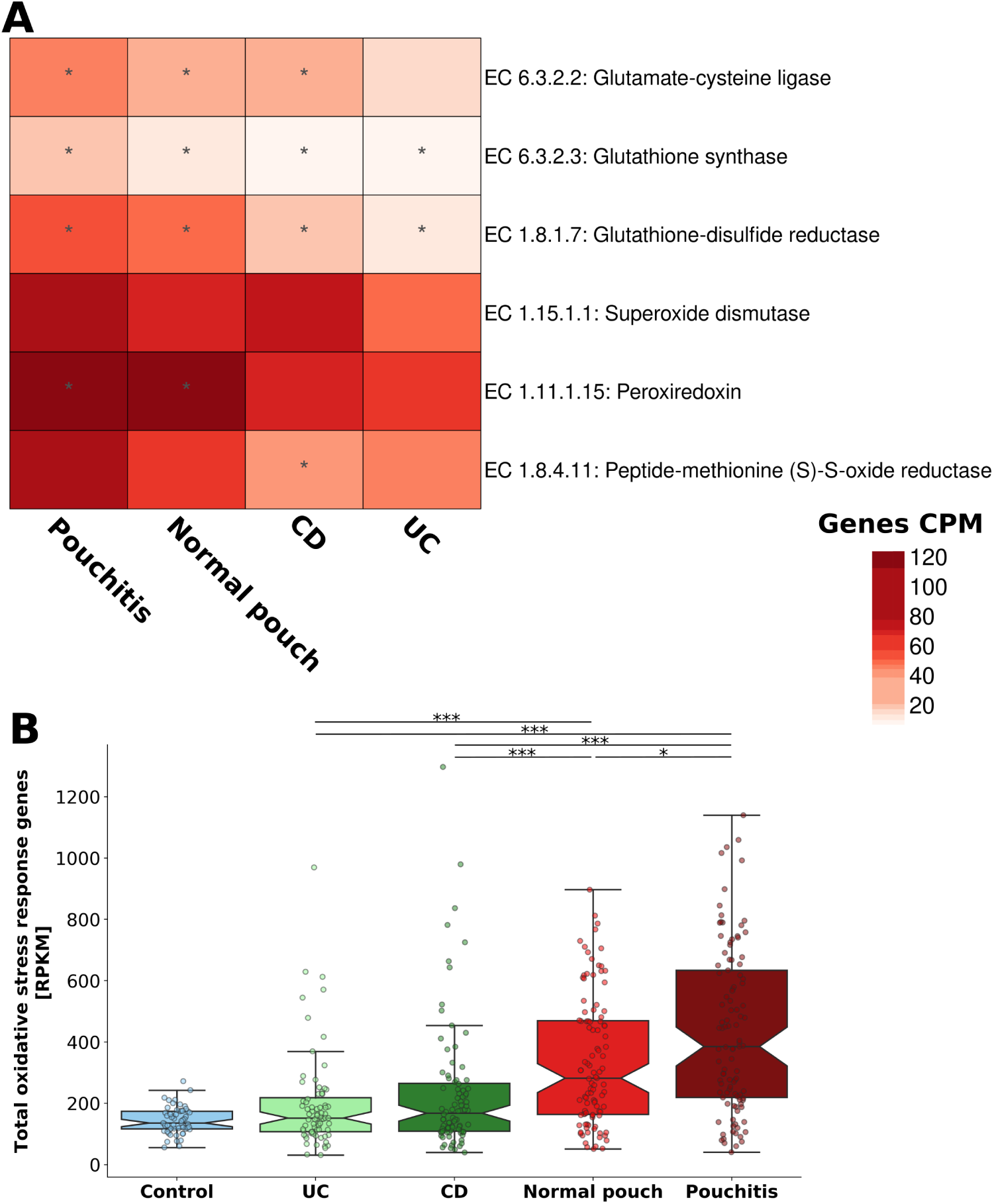
Total oxidative stress response in the microbiome. (**A**) Oxidative stress response enzymes (from Fig. 6) associated with IBD phenotypes. A generalized linear mixed-effects model was built for each gene, and IBD phenotypes were set as a predictor (see legend of Fig. 2). Asterisks indicate patient groups for which the association was significant (FDR < 0.2). (**B**) The combined contribution of these six oxidative stress response enzymes in metagenomes of patients with IBD and healthy subjects. *P < 0.05; **P < 0.01; ***P < 0.001; Kruskal-Wallis, applying Dunn’s multiple correction test. For the list of all pairwise P-values see Supplementary Table 2. Boxplot whiskers mark observations within the 1.5 interquartile range of the upper and lower quartiles.

A clear difference was observed between the highly contributing species to these anti-oxidative stress enzymes in pouch compared to non-pouch phenotypes. In the former, the vast majority consisted of facultative anaerobes such as *E. coli* and to a lesser degree *Klebsiella* and few species of *Streptococcus* and *Lactobacillus*. In the latter, *E. coli* contribution was less pronounced and replaced by obligate anaerobes such as *Bacteroides* species, several Clostridiales species, *A. muciniphila, Bifidobacterium longum* and *Prevotella copri* (Fig 6). A similar trend of increased facultative anaerobes abundance was previously observed in patients with CD or UC compared to healthy individuals [36]. Our results suggest that microbes that are more oxygen-tolerant and encode more oxygen-detoxifying enzymes have a fitness advantage and become more abundant in the high oxidative environment of the pouch.

### Classification models to distinguish between patients with a normal pouch and pouchitis based on species and functional biomarkers

We observed multiple differences in species and functional genes between patients with a normal pouch and those with pouchitis, yet no bacterial feature on its own was highly discriminative. We, therefore, attempted to use bacterial composition, metabolic pathways and enzymes profiles in order to build classifiers that will distinguish between these pouch phenotypes in the discovery cohort. In addition, we used the classifier built on our discovery cohort to classify samples with recurrent-acute pouchitis from the validation cohort in order to test if the model is able to correctly predict whether these patients will become normal pouch (disease improvement) or pouchitis (disease aggravation) in follow-up clinic visits. We used a machine learning algorithm of gradient boosting trees (GBT, see Materials and Methods) to train the classifier and evaluated its performance using five-fold cross-validation.

The GBT classifier based on the full set of species profile (230 features) achieved a mean area under the curve (AUC) of 0.787 ± 0.067 (Fig. 8A) for correctly classifying pouch samples into normal pouch or pouchitis. To improve the model’s predictive power, we used the feature importance scores that the algorithm assigns to each feature used during training, therefore features with the highest scores are the most informative for classification (Materials and Methods). A GBT classifier trained on the 40-top scoring features (species) showed improved performance with AUC of 0.818 ± 0.058 (Fig. 8A). Slightly improved outcomes were noticed when fecal calprotectin was incorporated into the model as a predictor (AUC=0.835 ± 0.055). Next, we built GBT models with metabolic pathways (MetaCyc) and microbial enzymes (ECs) as features. The pathway-based GBT model with a full set of 395 features achieved AUC of 0.76 ± 0.065 (Fig. 8B), while a model with the 60-top scoring features achieved AUC of 0.834 ± 0.056, a similar performance to the species based classifier. Importantly, GBT models trained on bacterial enzymes achieved better classification scores than species- or metabolic pathways-based models, despite having a larger number of features (1646) (Fig. 8C). The model with a full set of enzymes had an AUC of 0.802 ± 0.064, whereas, in a model with a reduced set of topmost 100 features, the AUC increased to 0.91 ± 0.046 (Fig. 8C). Fecal calprotectin did not notably improve the pathways and enzymes models. Furthermore, as the sole predictive feature, fecal calprotectin achieved a poor AUC of 0.625 ± 0.077 (Fig. 8D) in distinguishing normal pouch from pouchitis patients.

**Fig. 8.**
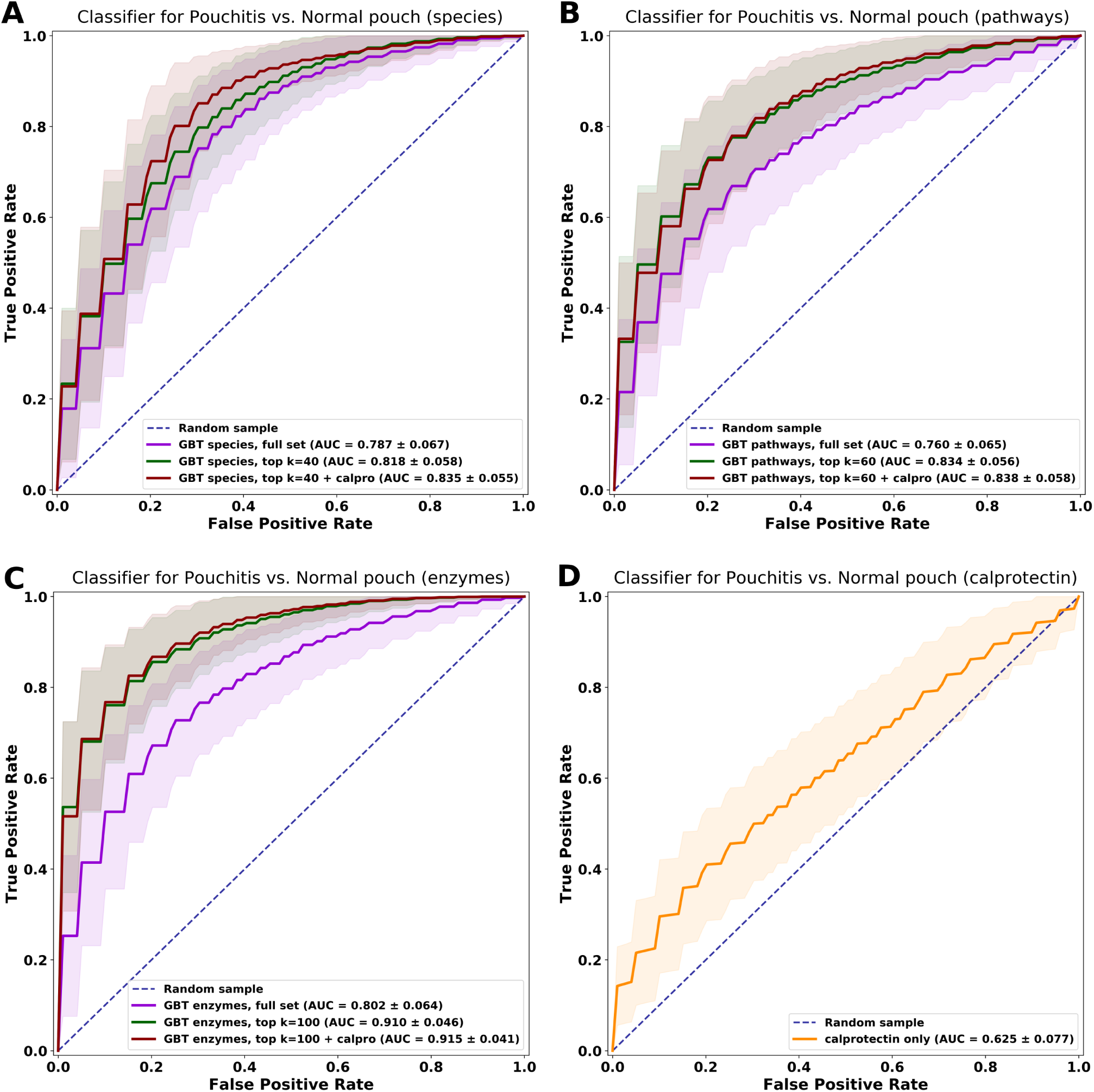
Classification models to distinguish between patients with a normal pouch and pouchitis. We trained and evaluated gradient boosting trees (GBT) classifiers on bacterial composition, metabolic pathways and enzymes profiles from the metagenomes of patients with a pouch (discovery cohort) by using five-fold cross-validation (randomly repeated 100 times). Area under the curve (AUC) measure (mean ± standard deviation) of each classifier performance is presented. (**A**) Species based classifier. (**B**) Metabolic pathways-based classifier. (**C**) Enzymes-based classifier. (**D**) Fecal calprotectin-based classifier. For each different classifier (species-, pathways- and enzymes-based, except for calprotectin only), three different models were built, with a full set of features (purple), reduced set with k=top scoring features and with a reduced set of features and fecal calprotectin as an additional predictor.

By analyzing the best predicting microbial features, we can gain new insights and identify potential biomarkers to disease pathogenesis. Reassuringly, among the highest-scoring features (species, pathways and enzymes) our models selected to best distinguish patients with pouchitis from a normal pouch, appeared many of the features mentioned above that differed significantly between these phenotypes and with potential clinical relevance to pouchitis pathogenesis (Supplementary Fig. 7, Supplementary Table 6).

Finally, we used the best species model (with 40 top scoring features) trained on the discovery cohort (n=208 samples), to test the model’s ability to correctly classify samples with recurrent-acute pouchitis phenotype from the validation cohort (n=42 samples) according to their future phenotype. We defined the true label of these samples according to the last recorded phenotype during clinic visit (future clinical observation compared to the current sample), when these patients’ phenotype changed either to a normal pouch or to pouchitis. Our species model (with or without calprotectin as an additional predictor) achieved an AUC of 0.778 (Supplementary Fig. 8A), but other performance metrics were better when including calprotectin: accuracy 76.2%, sensitivity: 88.9% and specificity: 53.3%. Notably, the sensitivity was high, the model correctly classified 24/27 recurrent-acute cases that later changed to pouchitis, however, the specificity was very low and only 8/15 that changed to normal pouch were classified correctly (Supplementary Fig. 8B). This implies that our classifier has a tendency to classify patients with recurrent-acute pouchitis more often into “pouchitis” rather than “normal pouch” and that recurrent-acute patients have a microbial profile more similar to patients with pouchitis.

In summary, given the complexity of the pouch microbiome, the challenge in classifying patients into normal pouch or pouchitis based on metagenomic biomarkers is not surprising. Nonetheless, functional genes had better classifying power than species composition, as the former incorporate enzymatic functions that are sometimes strain-specific (as in the *R. gnavus* example above) as well as those functions that can be encoded by many diverse species. It is possible to “predict” which patients with recurrent-acute pouchitis phenotype will develop into pouchitis or improve into a normal pouch, but this requires further validation in a larger cohort of such patients. Importantly, these analyses support our notion that not only does the pouch represent an unstable milieu, this milieu tends to be pro-inflammatory in nature.

## Conclusions

In this study, we explored bacterial functional dysbiosis in a well-phenotyped cohort of patients with a pouch. We identified several metabolic pathways and enzymes that were altered in patients with a pouch compared to patients with CD or UC and healthy controls. Most notably, the functional dysbiosis was more severe in patients with pouchitis, characterized by a shortage of genes for synthesis of butyrate and secondary bile acids. Moreover, patients with pouchitis harbored more facultative anaerobic bacteria encoding anti-oxidative stress enzymes, suggesting high oxidative stress during pouch inflammation. It appears that there is no excessive bacterial mucus utilization and degradation in the pouch, but rather a change in the mucolytic bacterial community composition, highlighted by an increased fraction of *R. gnavus* strains encoding fucosidases. We propose that the normal pouch and more so pouchitis are already dysbiotic (function- and species-wise) and based on multiple features, pouch phenotypes are more similar to CD. Our machine learning models emphasize the importance of integrating microbial-based biomarkers into clinical diagnostic tools. Our findings also delineate microbial and functional factors that underlie the pathogenesis of pouchitis and may suggest pathways such as butyrate that could be targeted for intervention in combination with dietary changes such as higher intake of fibers, to attenuate small intestinal inflammation, such as in pouchitis and CD.

## List of abbreviations

IBD: inflammatory bowel diseases
CD: Crohn’s disease
UC: ulcerative colitis
SCFA: short-chain fatty acid
CLDP: Crohn’s like disease of the pouch
EC: Enzyme Commission number
GH: glycoside hydrolases
GBT: gradient boosting trees
AUC: under the curve
DCA: deoxycholic acid
LCA: lithocholic acid
RPKM: reads per kilobase per million mapped reads
cpm: copies per million

## Declarations

### Ethics approval and consent to participate

The study was approved by the local institutional review board (0298–17) and the National Institutes of Health (NCT01266538). All patients signed informed consent before inclusion to the study.

### Consent for publication

Not applicable.

## Authors’ contributions

V.D., U.G. and I.D. conceived and designed the study; V.D. developed the bioinformatic analysis and machine learning pipelines and analyzed the metagenomic data; L.R. analyzed the metagenomic data; K.R., L.G., K.Y., K.Z. and I.D. collected, analyzed and provided the clinical data; N.W. and I.D. enrolled and examined the patients; L.G. provided and analyzed the dietary information. V.D., L.R., U.G. and I.D. wrote the paper. All authors read, discussed, and approved the final manuscript.

## Acknowledgments

The authors wish to thank the members of the RMC IBD Center and the Comprehensive Pouch Clinic for their help and stimulating discussion of the project. The authors wish to thank Stefan Green of the University of Illinois at Chicago, for his continued expert help in the metagenomic sequencing.

## Supplementary figures

**Supplementary Fig. 1.**
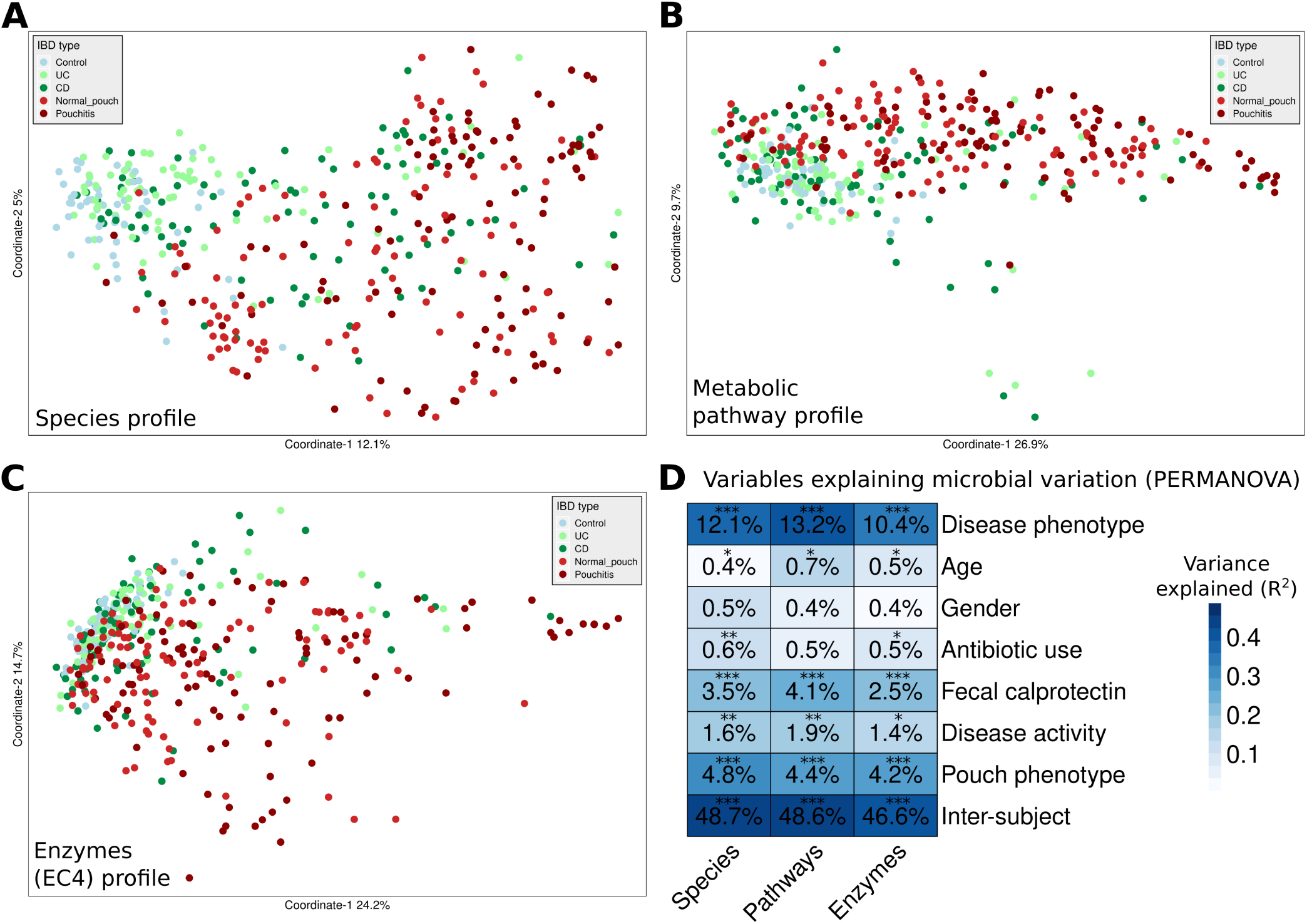
The microbiome variation (principal coordinates analysis, PCoA) based on Bray-Curtis dissimilarity for (**A**) species, (**B**) pathways and (**C**) enzymes compositional profiles of the fecal metagenomes. (**D**) Variables explaining the variation in microbiome species, pathways and enzymes, analyzed with PERMANOVA. Percentages indicate variance explained by each variable (R^2^ values) independently of the other variables. *P < 0.05; **P < 0.01; ***P < 0.001.

**Supplementary Fig. 2.**
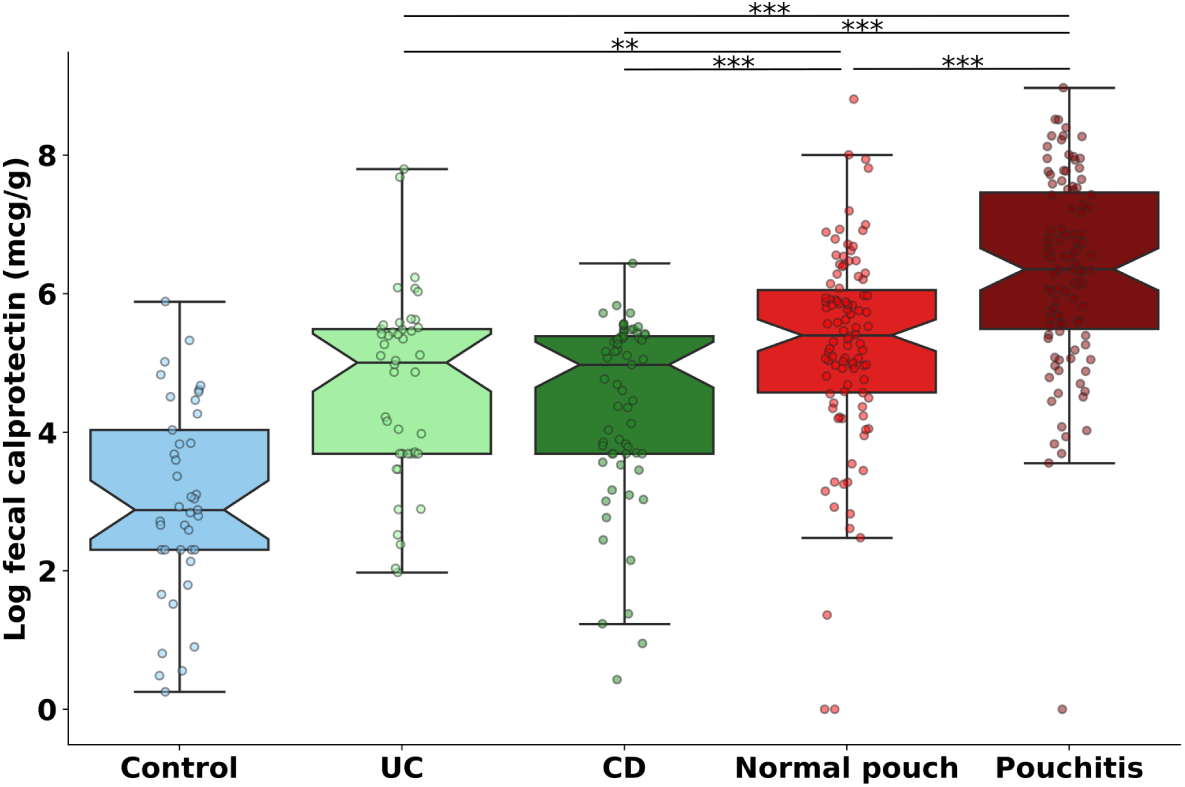
Fecal calprotectin levels (log-transformed) in patients with IBD and healthy controls. *P < 0.05; **P < 0.01; ***P < 0.001; Kruskal-Wallis, applying Dunn’s multiple correction test. For the list of all pairwise P-values see Supplementary Table 2. Boxplot whiskers mark observations within the 1.5 interquartile range of the upper and lower quartiles. Fecal calprotectin measurements were available for 358 out of 428 samples (see Supplementary Table 1).

**Supplementary Fig. 3.**
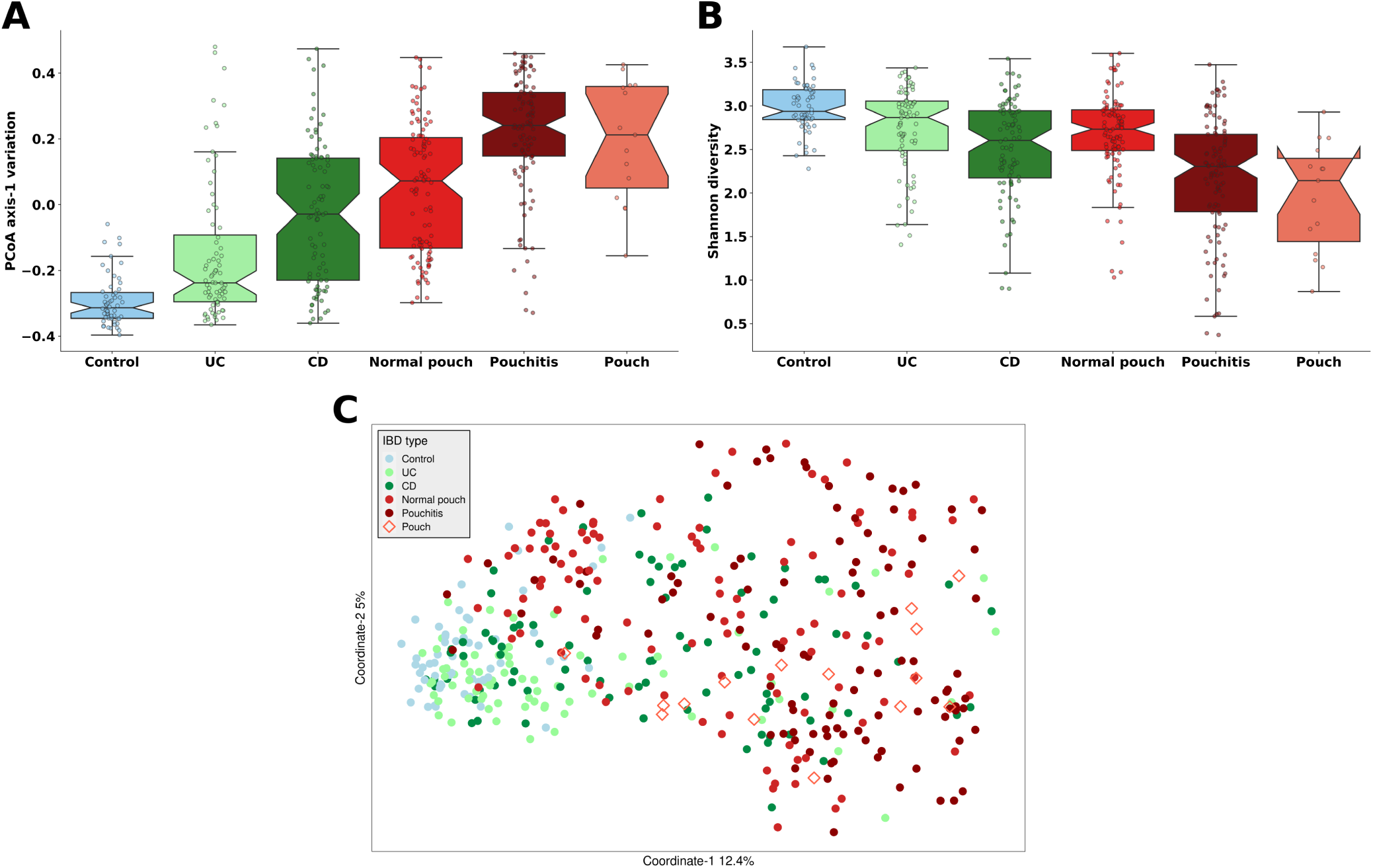
Comparison of 15 publicly available pouch metagenomes from an American cohort (^31^Sinha et al. 2020) to the rest of the pouch and non-pouch metagenomes analysed in the current study. (**A**) The microbiome variation according to the first axes of principal coordinates analysis (PCoA) of species profiles, based on Bray-Curtis dissimilarity of the fecal metagenomes. (**B**) Shannon diversity in the microbiome based on species composition. (**C**) PCoA analysis of species profiles based on Bray-Curtis dissimilarity of the fecal metagenomes. The samples labeled as “Pouch” are the pouch metagenomes from ^31^Sinha et al. 2020.

**Supplementary Fig. 4.**
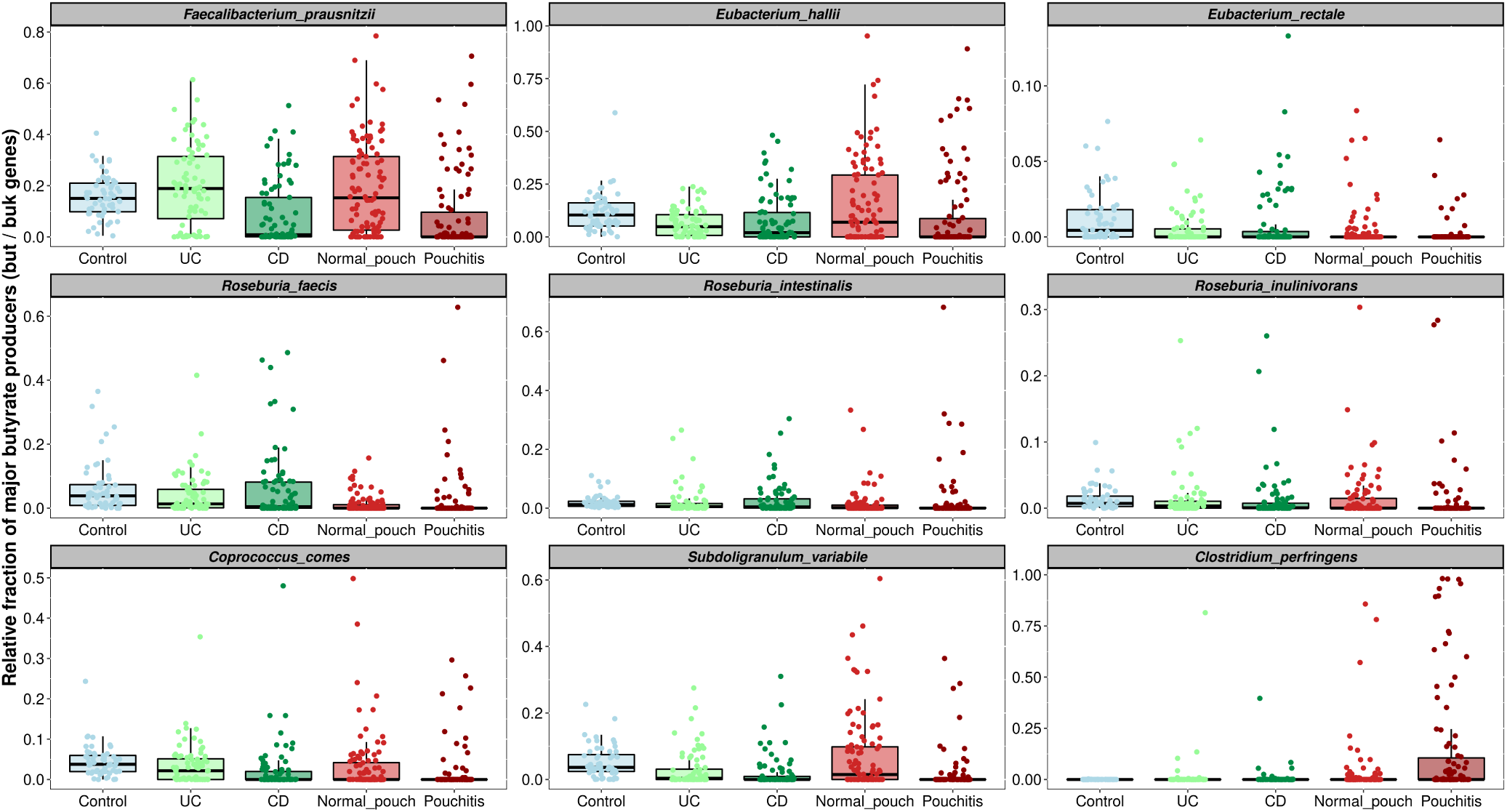
The Major butyrate producers in the fecal metagenomes of patients with IBD and healthy controls. The relative fraction and taxonomic affiliation of the most abundant butyrate-producing bacteria based on the similarity of the identified *but* (butyryl-CoA:acetate CoA transferase) and *buk* (butyrate kinase) genes to known butyrate producers. *Coprococcus comes, Subdoligranulum variabile* and *Clostridium perfringens* were quantified based on *but*, and the rest of the species based on *but*. For a complete list of all the identified butyrate producers, see Supplementary Table 4

**Supplementary Fig. 5.**
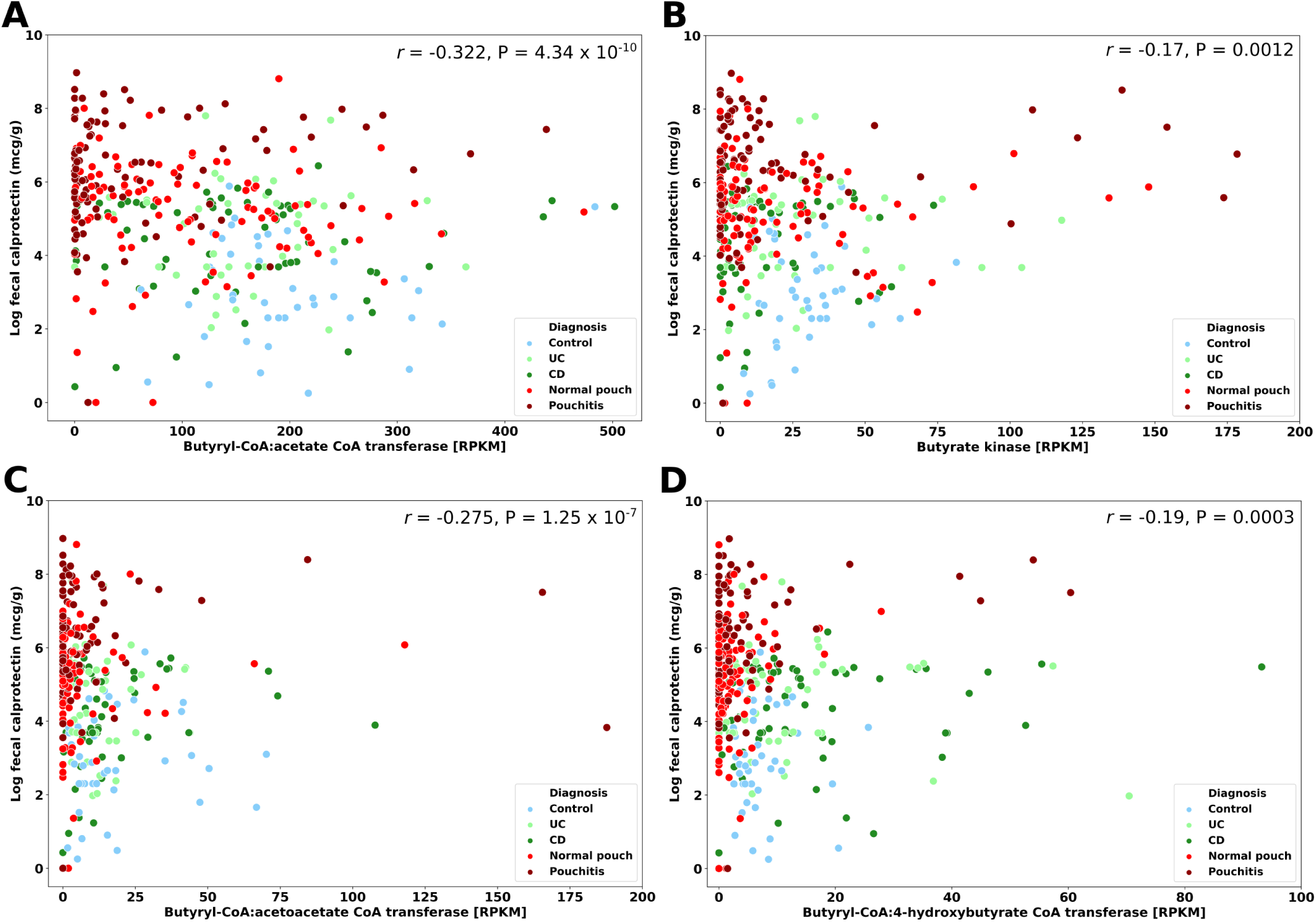
Spearman correlation between the abundance of butyrate synthesis genes (*but, buk, ato* and *4hbt*) and fecal calprotectin levels in patients with IBD and healthy controls. Fecal calprotectin measurements were available for 358 out of 428 samples (see Supplementary Table 1).

**Supplementary Fig. 6.**
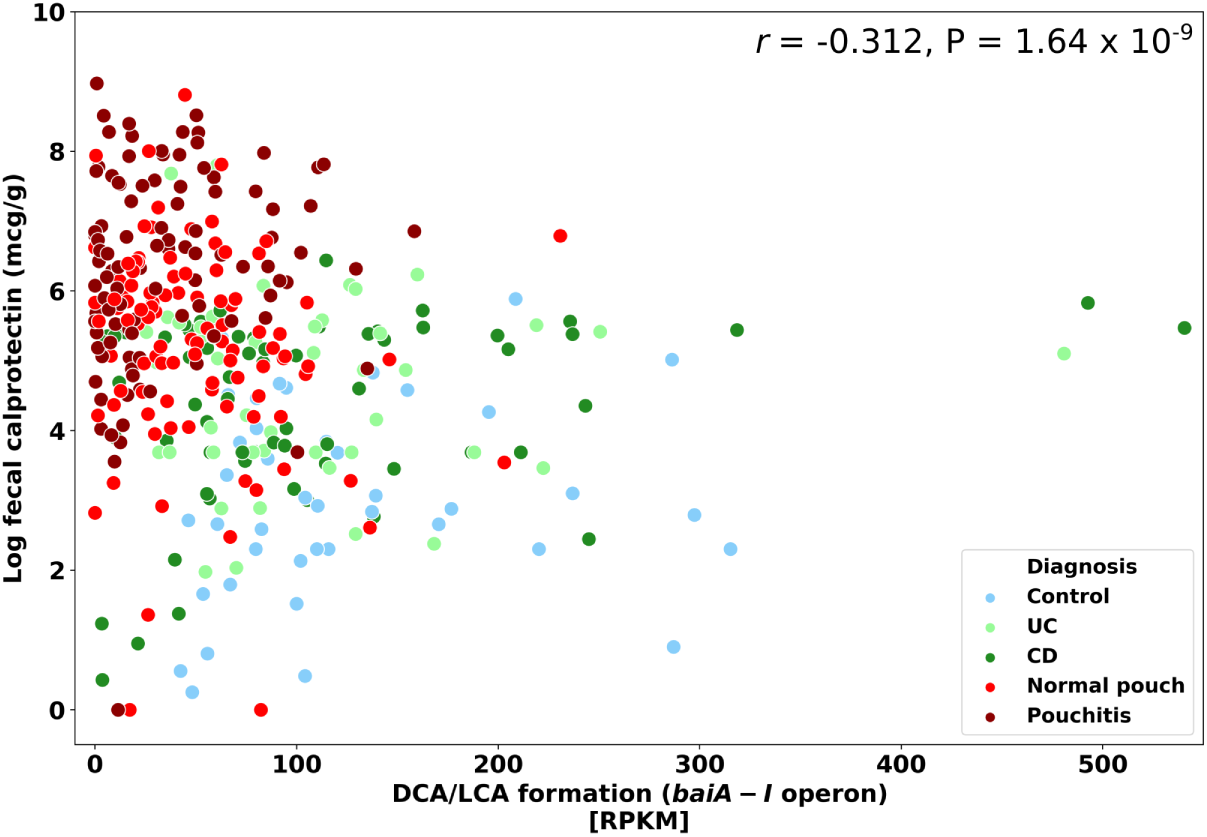
Spearman correlation between the abundance of secondary bile acids formation genes, *bai* (*baiA-I* operon) and fecal calprotectin levels in patients with IBD and healthy controls. Fecal calprotectin measurements were available for 358 out of 428 samples (see Supplementary Table 1).

**Supplementary Fig. 7.**
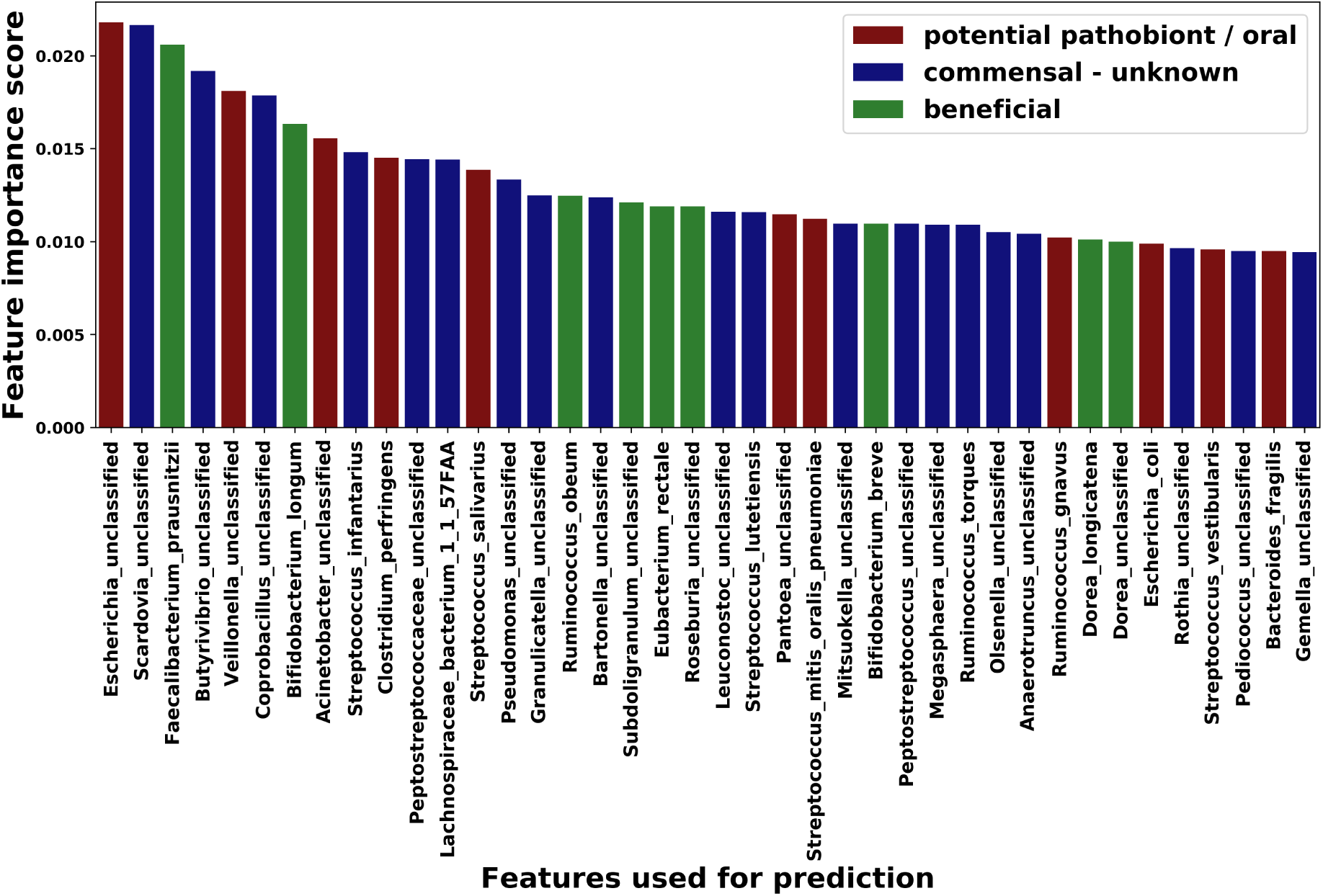
Highest scoring species features (most informative for classification) used in the classification model (of Fig. 8A) to distinguish between patients with a normal pouch and pouchitis. The average importance scores for the top 40 features are displayed, divided into “potentially beneficial bacteria”, “potential pathobiont / oral bacteria” and “unknown commensals” based on recent literature consensus. For the full list of scores for each species, see Supplementary Table 6.

**Supplementary Fig. 8.**
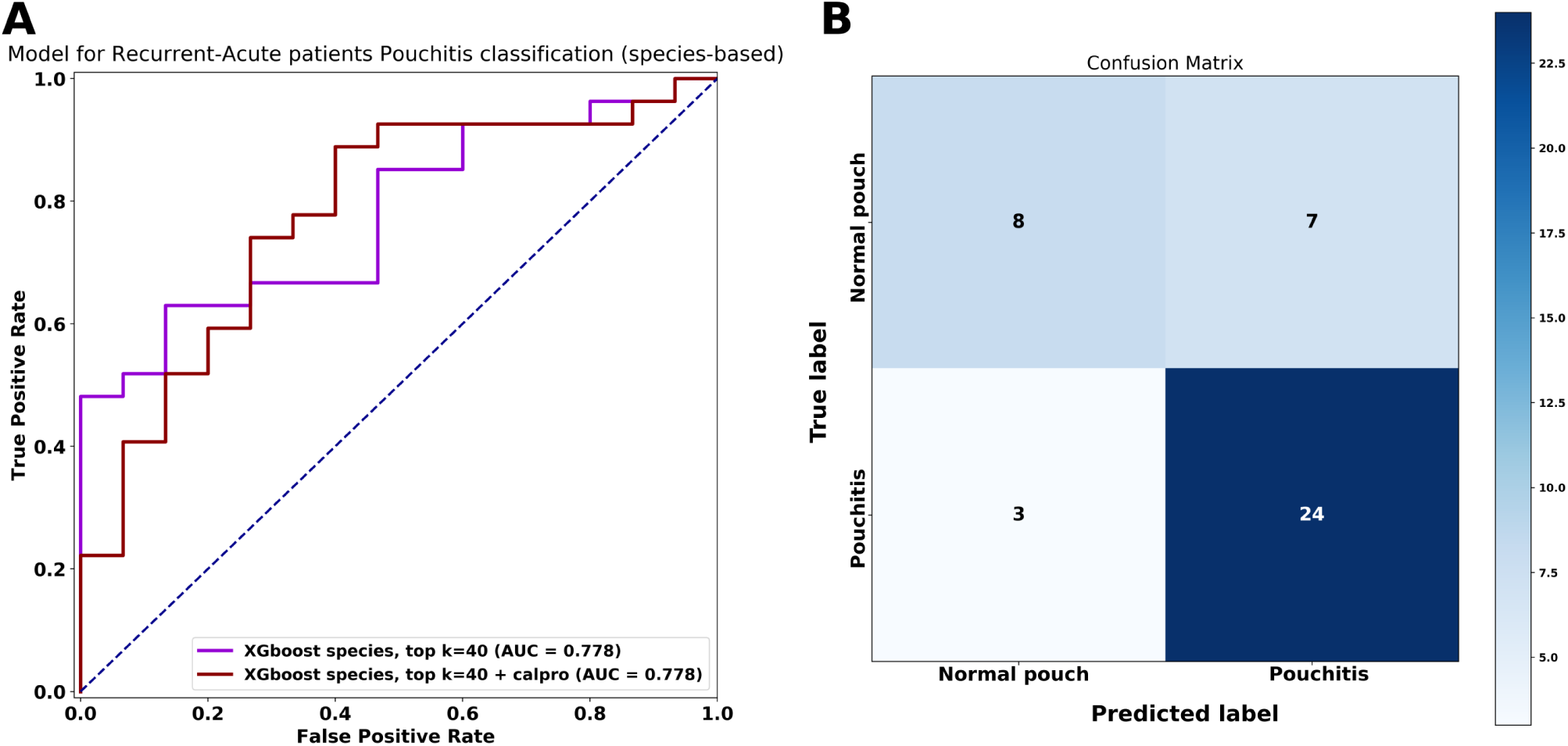
Testing the model performance (best species-based with k=40 top features) trained on the discovery cohort (n=208 samples) in classifying samples from the validation cohort (n=42 samples) with recurrent-acute pouchitis phenotype, into a normal pouch or pouchitis according to the last observed phenotype (by future clinical observation). (**A**) Area under the curve (AUC) measure of two classifiers (with or without calprotectin as an additional predictor) (**B**) Confusion matrix evaluation of the classifier (with calprotectin), comparing the number of correctly and incorrectly classified samples. 24/27 recurrent-acute samples that later changed to pouchitis and 8/15 that later changed to a normal pouch, were classified correctly, respectively.

## Supplementary methods

### Pouch Disease Behavior (pouch phenotype)

Pouch disease behavior was defined as normal, acute/recurrent-acute pouchitis, chronic pouchitis, or CLDP, as previously defined [1]. Briefly, a normal pouch was defined as no symptoms of pouchitis during the past 2 years and no antibiotic therapy. Acute/recurrent-acute pouchitis was defined as a flare of pouchitis responding to a short course (usually 2 weeks) of antibiotic therapy, or up to 4 flares/year, respectively. Chronic pouchitis was defined as >4 pouchitis flares/year or administration of antibiotics or IBD-specific anti-inflammatory therapy for more than 1 month. CLDP was defined as having pouch-perianal disease, pouch strictures, or long segments of proximal small intestinal inflammation. Patients undergoing pouch surgery due to familial adenomatous polyposis (FAP) were recruited as well and had a normal pouch throughout follow-up.

### Dietary information based on food frequency questionnaires

Intake of food groups and nutrients were assessed using Food Frequency Questionnaires (FFQs) adapted to the Israeli population and administered in “MABAT” Israeli nutrition and public health governmental study [2]. For each of the 106 food items, a commonly used portion size was specified. Patients were asked how often on average they consumed each food item and daily intake of nutrients was calculated by multiplying the frequency of each food item by the nutrient content based on the Israeli TZAMERET food database and the United States Department of Agriculture (USDA) food database [3].

